# Addressing Personal Protective Equipment (PPE) Decontamination: Methylene Blue and Light Inactivates SARS-CoV-2 on N95 Respirators and Masks with Maintenance of Integrity and Fit

**DOI:** 10.1101/2020.12.11.20236919

**Authors:** Thomas S. Lendvay, James Chen, Brian H. Harcourt, Florine E.M. Scholte, F. Selcen Kilinc-Balci, Ying Ling Lin, Molly M. Lamb, Larry F. Chu, Amy Price, David Evans, Yi-Chan Lin, Christopher N. Mores, Jaya Sahni, Kareem B. Kabra, Eric Haubruge, Etienne Thiry, Belinda Heyne, Jan Laperre, Sarah Simmons, Jan M. Davies, Yi Cui, Thor Wagner, Tanner Clark, Sarah J. Smit, Rod Parker, Thomas Gallagher, Emily Timm, Louisa F. Ludwig-Begall, Nicolas Macia, Cyrus Mackie, Karen Hope, Ken Page, Susan Reader, Peter Faris, Olivier Jolois, Alpa Patel, Jean-Luc Lemyre, Vanessa Molloy-Simard, Kamonthip Homdayjanakul, Sarah R. Tritsch, Constance Wielick, Mark Mayo, Rebecca Malott, Jean-Francois Willaert, Hans Nauwynck, Lorène Dams, Simon De Jaeger, Lei Liao, Mervin Zhao, Steven Chu, John M. Conly, May C. Chu

## Abstract

**Background:** The coronavirus disease 2019 (COVID-19) pandemic has resulted in severe shortages of personal protective equipment (PPE) necessary to protect front-line healthcare personnel. These shortages underscore the urgent need for simple, efficient, and inexpensive methods to decontaminate SARS-CoV-2-exposed PPE enabling safe reuse of masks and respirators. Efficient decontamination must be available not only in low-resourced settings, but also in well-resourced settings affected by PPE shortages. Methylene blue (MB) photochemical treatment, hitherto with many clinical applications including those used to inactivate virus in plasma, presents a novel approach for widely applicable PPE decontamination. Dry heat (DH) treatment is another potential low-cost decontamination method.

**Methods:** MB and light (MBL) and DH treatments were used to inactivate coronavirus on respirator and mask material. We tested three N95 filtering facepiece respirators (FFRs), two medical masks (MMs), and one cloth community mask (CM). FFR/MM/CM materials were inoculated with SARS-CoV-2 (a *Betacoronavirus*), murine hepatitis virus (MHV) (a *Betacoronavirus*), or porcine respiratory coronavirus (PRCV) (an *Alphacoronavirus*), and treated with 10 µM MB followed by 50,000 lux of broad-spectrum light or 12,500 lux of red light for 30 minutes, or with 75°C DH for 60 minutes. In parallel, we tested respirator and mask integrity using several standard methods and compared to the FDA-authorized vaporized hydrogen peroxide plus ozone (VHP+O_3_) decontamination method. Intact FFRs/MMs/CM were subjected to five cycles of decontamination (5CD) to assess integrity using International Standardization Organization (ISO), American Society for Testing and Materials (ASTM) International, National Institute for Occupational Safety and Health (NIOSH), and Occupational Safety and Health Administration (OSHA) test methods.

**Findings:** Overall, MBL robustly and consistently inactivated all three coronaviruses with at least a 4-log reduction. DH yielded similar results, with the exception of MHV, which was only reduced by 2-log after treatment. FFR/MM integrity was maintained for 5 cycles of MBL or DH treatment, whereas one FFR failed after 5 cycles of VHP+O_3_. Baseline performance for the CM was variable, but reduction of integrity was minimal.

**Interpretation:** Methylene blue with light and DH treatment decontaminated masks and respirators by inactivating three tested coronaviruses without compromising integrity through 5CD. MBL decontamination of masks is effective, low-cost and does not require specialized equipment, making it applicable in all-resource settings. These attractive features support the utilization and continued development of this novel PPE decontamination method.

## INTRODUCTION

The coronavirus disease 2019 (COVID-19) pandemic caused by SARS-CoV-2 has resulted in critical personal protective equipment (PPE) shortages, especially filtering facepiece respirators (FFRs, also known as N95 respirators) and medical masks (MMs). N95 FFRs are respiratory protective devices which are designed to filter at least 95% of airborne particles. A MM is a medical device covering the mouth, nose and chin ensuring a barrier that limits exposure of an infective agent between healthcare personnel (HCP) and the patient. MMs are used by HCPs to prevent large respiratory droplets and bodily fluid splashes from reaching the mouth and the nose of the wearer and help reduce and/or control at the source the spread of large respiratory droplets from the person wearing the mask. MMs include surgical and procedure masks and are not considered respiratory protective devices. N95 FFRs tightly fit the face while MMs are designed as loose-fitting devices. Although designed for single short-term usage, FFRs and MMs potentially contaminated by SARS-CoV-2 are being re-used on an emergency basis due to short supply during the current pandemic. These shortages have necessitated the rapid development and deployment of various disinfection techniques and equipment, and have led to the World Health Organization (WHO) issuing interim guidance on rational PPE use (1), including methods for decontamination (mostly for FFRs) (2). The U.S. Food and Drug Administration (FDA) granted Emergency Use Authorization (EUA) of hydrogen peroxide and steam sterilization systems to decontaminate FFRs/MMs intended for reuse (3). However, these technologies remain less available in low-resource settings, where the majority of frontline HCPs have inadequate protection.

Photochemical treatment represents a disinfection method that uses a photosensitive drug, referred to as a photosensitizer, which when combined with visible light, generates singlet oxygen from ambient molecular oxygen in the air. Singlet oxygen inactivates viruses by damaging viral nucleic acids and viral membranes (4,5). One such photosensitizer is methylene blue (MB), which, in addition to its primary FDA-approved medical indication to treat methemoglobinemia, has been used as a sterilization method for human plasma transfusions when combined with light, used for inactivating SARS-CoV-2 and many other microorganisms (6-8). See Supplemental material for additional background information.

The aim of the “Development of Methods for Masks and N95 Decontamination” (DeMaND) study was to evaluate methods that inactivate SARS-CoV-2 on respirators and masks, which can be applied anywhere, at low cost and without complicated procedures. We examined if methylene blue plus light (MBL) or dry heat (DH) treatments could effectively decontaminate commonly used FFRs, MMs, or a community mask (CM) while maintaining mask integrity (filtration, breathability, fluid resistance and fit) after multiple cycles of decontamination. This study leveraged four virology laboratories and six PPE integrity testing sites to examine: (a) MBL and DH virucidal effect using two isolates of SARS-CoV-2 and two other coronaviruses requiring a lower level of biocontainment, and (b) the impact of five cycles of decontamination (5CD) on the integrity of treated FFRs/MMs and a CM [FIGURE 1].

**FIGURE 1.**
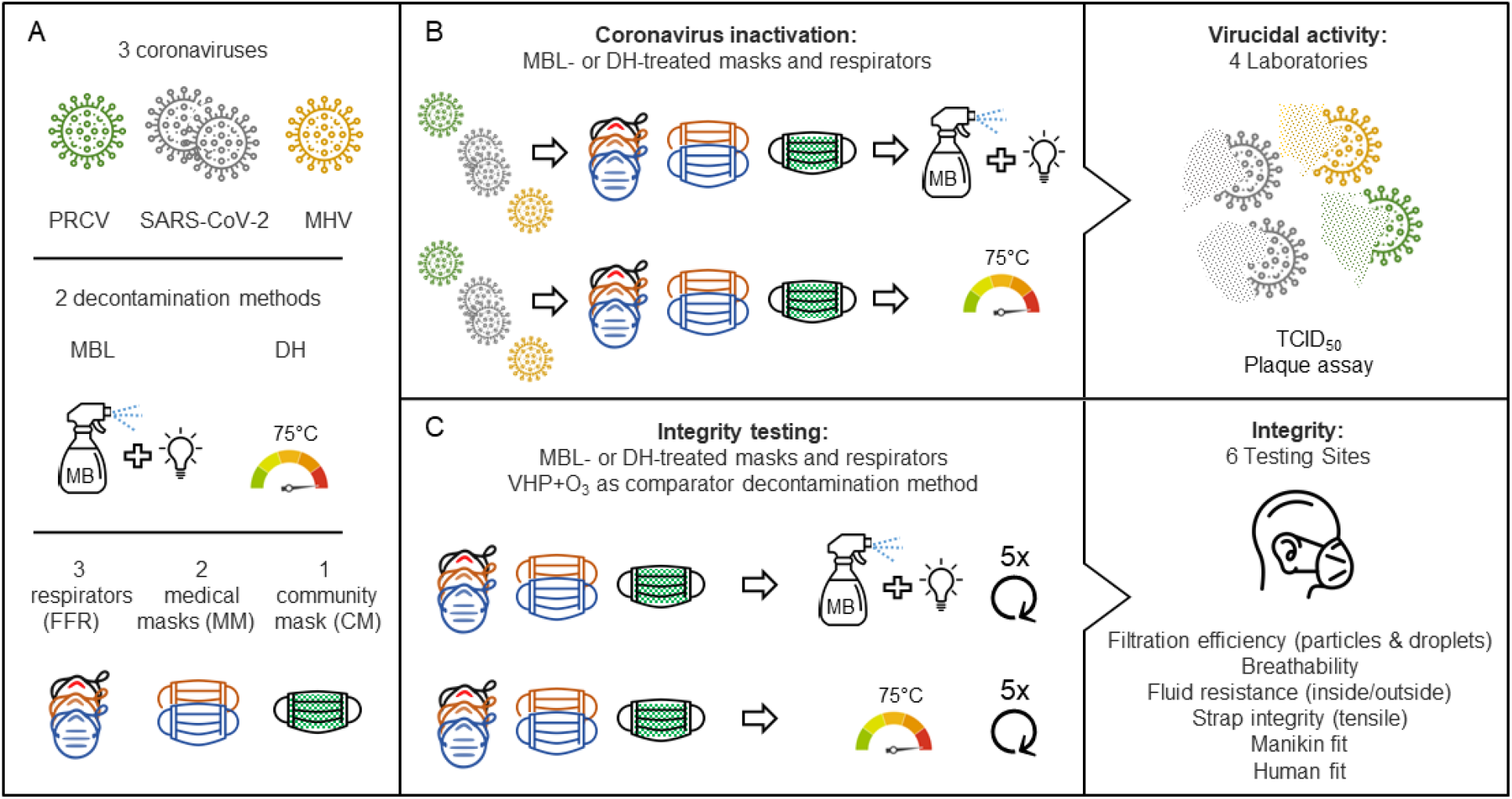
Graphical representation of the DeMaND study outline. **(A)** Overview of the coronaviruses, decontamination methods and masks used. **(B)** FFRs, MMs andr a CM were inoculated with virus and treated with either MB and light (MBL) or dry heat (DH). Remaining infectious virus was quantified using TCID_50_ or plaque assays. **(C)** In parallel with the virucidal testing of MBL and DH treatments, intact FFRs/MMs/CMs were subjected to 5 cycles of decontamination before mask integrity/fit was tested using the indicated methods. Abbreviations: PRCV = porcine respiratory coronavirus, SARS-CoV-2 = severe acute respiratory syndrome coronavirus 2, MHV = murine hepatitis virus, MBL = methylene blue + light, DH = dry heat (75°C), VHP+O_3_ = vaporized hydrogen peroxide plus ozone. See Supplemental Table S1 for the respirator and mask decontamination and testing matrix.

The number of decontamination cycles was selected based on the Centers for Disease Control and Prevention (CDC)’s recommended maximum number of donnings as part of crisis capacity strategies based on the current literature (9). The FFRs/MMs were selected based on popularity and availability during recent outbreak response and to account for physical differences such as cup shapes, material and structure. For the integrity testing, we compared MBL and DH to the FDA-authorized vaporized hydrogen peroxide plus ozone system (VHP+O_3_) (2).

## RESULTS

### Methylene blue and light (MBL) inactivates coronaviruses in tissue culture plates

To confirm that MBL is able to inactivate coronavirus, varying concentrations of MB were mixed with porcine respiratory coronavirus (PRCV) and exposed to light (12,500 lux of red light). Treatment with 0.1 µM MB plus light resulted in a 6-log reduction in PRCV titers. In the absence of additional light, a higher MB dose (1 µM) still resulted in a 6-log reduction [FIGURE 2A].

**FIGURE 2.**
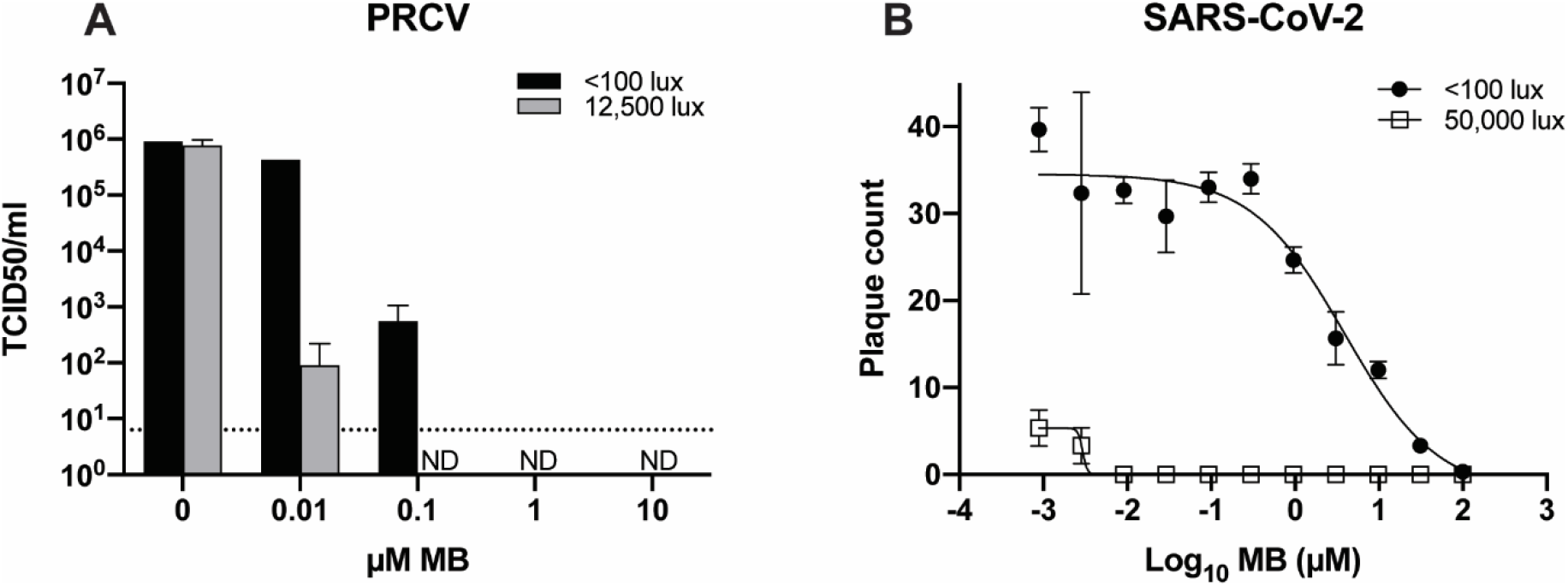
Inactivation of PRCV and SARS-CoV-2 using methylene blue and light (MBL). **(A)** To determine the efficacy of different MB concentrations, serial dilutions of MB were added to wells of a 48-well plate containing 10 µL PRCV (2×10^7^ TCID_50_/ml). Technical triplicates were performed using three separate plates. Plates were either exposed to a red-light source (12,500 lux) for 30 minutes or were protected from light (<100 lux). Dotted line indicates limit of detection. **(B)** Serial dilutions of MB were added to wells of a 12-well plate containing ∼50 PFU of SARS-CoV-2 in MEM plus 15% FCS. Plates were either exposed to 50,000 lux of broad-spectrum light for 45 min or were protected from light (<100 lux). Viral titers were determined using three replicate samples.

Next, to confirm the ability of MBL to specifically reduce SARS-CoV-2 infectivity, varying concentrations of MB were mixed with SARS-CoV-2 and exposed to high intensity broad-spectrum light (50,000 lux). MB inhibited SARS-CoV-2 infectivity with a dose-dependent effect, with or without exposure to high intensity light. This virucidal effect was strikingly enhanced in the presence of a strong light source. The half-maximal effective concentration (EC_50_) of MB was 3.1 µM when exposed to <100 lux of light, but 1.8 nM with the addition of high intensity light, a 1722-fold effect [FIGURE 2B].

### MBL and DH inactivate coronaviruses on FFR/MM material

To examine the dose required to inactivate respirators/masks contaminated with SARS-CoV-2 or murine hepatitis virus (MHV), we tested a representative generic facemask and N95 respirator (Type II generic facemask (FW) and 3M 1870+ N95 respirator (R3), respectively). These masks were cut into coupons, inoculated with virus, and treated with 1 or 10 μM MB and light for the indicated time periods [FIGURE 3A]. All viruses displayed sensitivity to MBL treatment. Using 10 µM MB with light resulted in complete inactivation of SARS-CoV-2 and MHV on both FFR and MM material after 5 minutes of treatment: a 3-4 log reduction of SARS-CoV-2 and >5 log reduction of MHV titers. Using a lower dose (1 µM), complete inactivation was observed after 30 minutes of light exposure, though a 2-4 log reduction of viral titers was already observed after 5 minutes. We also observed that MB treatment in the absence of additional light resulted in substantial reductions of viral titers.

**FIGURE 3.**
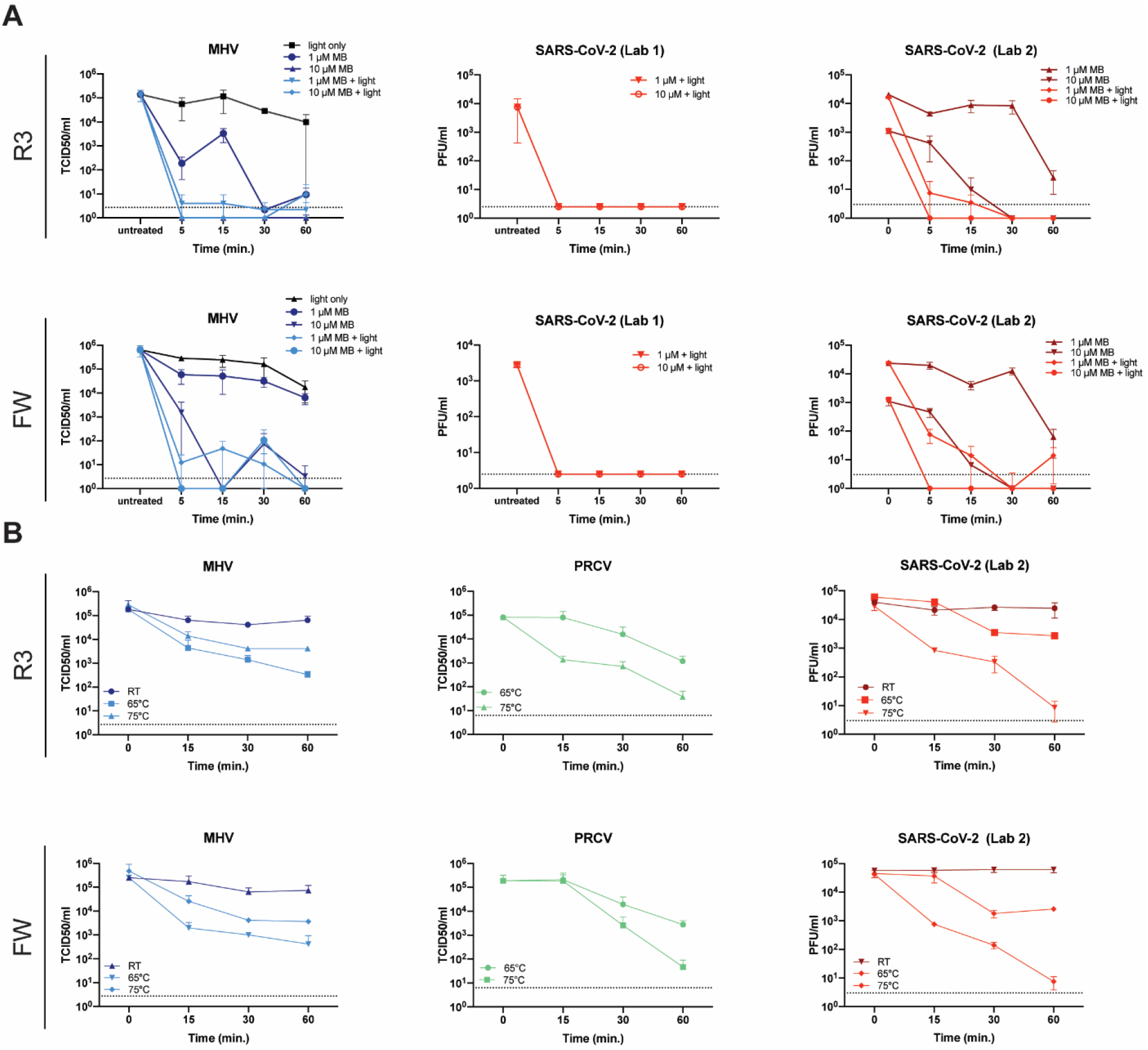
MBL and DH inactivate MHV, SARS-CoV-2 and PRCV on FFR/MM material. **(A)** Effect of methylene blue and light treatment on SARS-CoV-2 and MHV titers. A 10 µl aliquot of SARS-CoV-2 or MHV was applied to coupons derived from a FFR (R3) or MM (FW) and left to dry for 20 min. Subsequently 10 or 30 µl of MB was added to the virus spot on each coupon at the indicated concentrations. The samples were exposed to light (50,000 lux) for the indicated time periods or left in the biosafety cabinet with the lights off. Each virus titer was measured using 2-6 replicate samples by plaque assay or median tissue culture infection dose (TCID_50_). Data are represented as mean +/-SD. **(B)** Effect of DH treatment on SARS-CoV-2, PRCV and MHV titers. A 10 µl aliquot of SARS-CoV-2 or MHV was applied to coupons derived from a FFR (R3) or MM (FW) and left to dry for 20 min. Subsequently, the samples were incubated at 65°C or 75°C for the indicated time periods. Alternatively, 100 ul of PRCV was injected under the outer layer of intact FFRs or MMs and allowed to dry for 30 minutes before exposure to DH. Each virus titer was measured using 2-6 replicate samples by plaque assay or TCID_50_. Data are represented as mean +/-SD. FW= Type II generic face mask. R3= 3M panel respirator (1870+). Dotted line indicates limit of detection.

In parallel, coupons inoculated with SARS-CoV-2 or MHV, or complete masks inoculated with PRCV, were treated with 65°C or 75°C dry heat (DH) for up to 60 minutes. Although all viruses displayed a degree of temperature sensitivity, 60 minutes exposure did not result in complete inactivation [FIGURE 3B].

To ensure that MBL or DH can be utilized to efficiently decontaminate a wide variety of masks, we tested 3 more masks, including one additional MM (Type IIR ASTM F2100 Level 2 Halyard (FH)), and two additional FFRs (duckbill FFR from Halyard (RH) and half-sphere 1860 from 3M (RM)). Coupons or intact masks were inoculated as described, treated with 10 µM MB and exposed to light for 30 minutes [FIGURE 4A-D], conditions which demonstrated robust inactivation in the previous experiments. We observed complete inactivation (up to 4 logs) of SARS-CoV-2 for all masks and respirators tested. Treatment with 10 µM MB without exposure to bright light resulted in substantial virus reduction [FIGURE 4B]. For MHV, no virus was detectable for FH, R3, RH, and RM, signifying a 4-5 log reduction. A low level of virus was detectable in one replicate for FW [FIGURE 4A]. For PRCV, which was injected underneath the outer mask layer, a >5-log virus reduction was observed after MB treatment of FH, FW, R3 and RH masks. In contrast, only an approximate 3-log reduction was observed for RM masks [FIGURE 4D].

**FIGURE 4.**
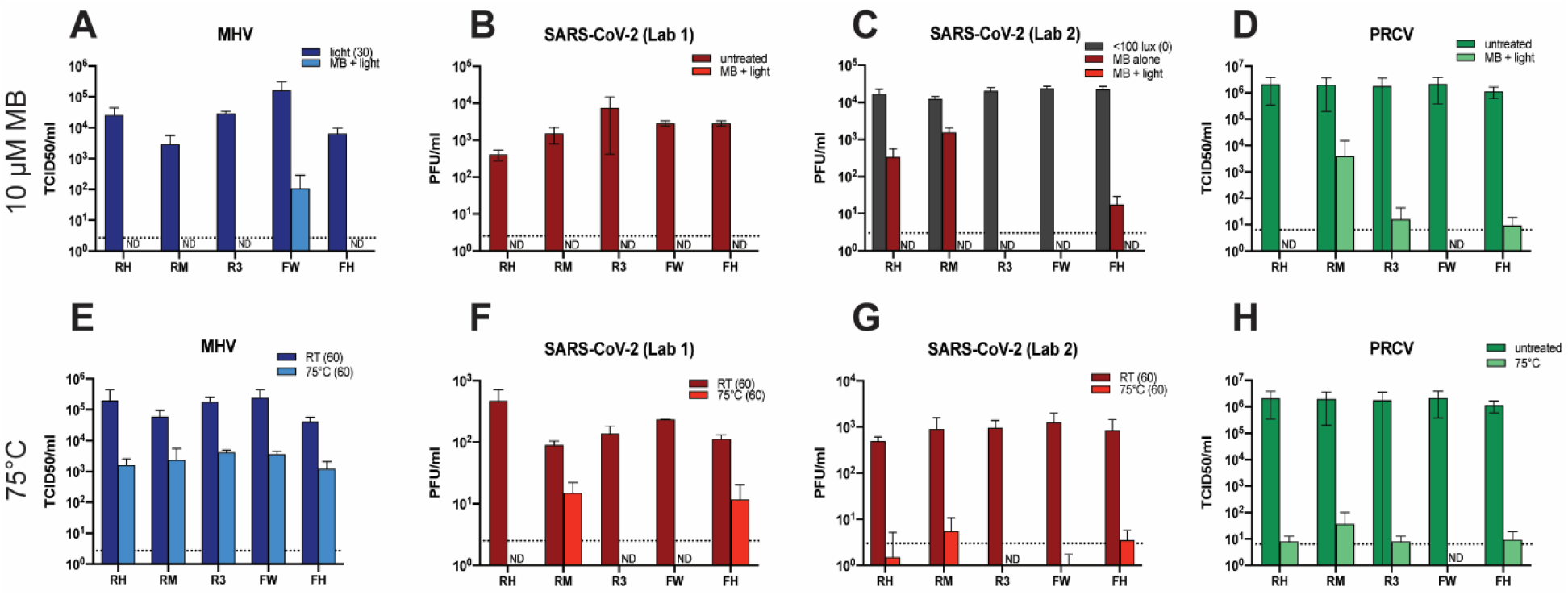
Effect of MBL treatment on MHV, PRCV and SARS-CoV-2 titers on the surfaces of three FFRs (R3, RH, RM) and two MMs (FH, FW). **(A-C)** A 10 µl aliquot of SARS-CoV-2 or MHV was applied to coupons of the indicated masks and dried for 20 min. 10-30 µl of 10 µM MB was added to each coupon, depending on coupon size, and the samples were treated with light (50,000 lux) or protected from light. **(D)** 100 ul of PRCV was injected under the outer layer of intact FFRs or MMs and allowed to dry for 30 minutes. Subsequently the FFRs and MMs were sprayed with 10 µM MB and dried for 30 minutes in the dark before exposure to 12,500 lux of red light. **(E-H)** Effect of 75°C DH treatment on MHV, PRCV and SARS-CoV-2 titers on the surface of **three FFRs (R3, RH, RM) and two MMs (FH, FW)**. A 10 µl aliquot of SARS-CoV-2 or MHV was applied to coupons of the indicated masks and dried for 20 min. **(H)** 100 ul of PRCV was injected under the outer layer of intact FFRs/MMs and dried for 30 minutes. For PRCV, MBL and DH were tested simultaneously and share the same control data. Coupons and intact respirators/masks were incubated at 75°C for 60 minutes. Each virus titer was determined using 2-6 replicate samples by plaque assay or TCID_50_. Data is represented as mean +/-SD. ND = Not detected. FH= Type IIR ASTM F2100 Level 2 Halyard face mask. FW= Type II EN 14683 generic face mask. R3= 3M panel respirator (1870+). RH= Halyard duckbill respirator (Fluidshield-46727). RM= 3M half-sphere respirator (1860). Dotted line indicates limit of detection.

To assess DH inactivation of these additional 3 masks, coupons or intact masks were inoculated with virus and exposed to 75°C for 60 minutes. PRCV was generally effectively inactivated by DH treatment, resulting in >5 logs of virus reduction [FIGURE 4H]. FH and RM coupons inoculated with SARS-CoV-2 were not fully inactivated [FIGURE 4F,G]. MHV titers were only modestly reduced (∼2 logs) across all mask types [FIGURE 4E]. Total log reductions for both MBL and DH conditions by mask type are listed in [TABLE 1].

**TABLE 1.** Log reduction and percent inactivation of coronaviruses treated with MBL or DH. Virus Reduction by 10 μM Methylene Blue vs. 75°C Dry Heat

| Condition | Mask type | MHV |  | PRCV |  | SARS-CoV-2 (Lab 1) |  | SARS-CoV-2 (Lab 2) |  | SARS-CoV-2 avg. |  | Overall avg. |  |
| --- | --- | --- | --- | --- | --- | --- | --- | --- | --- | --- | --- | --- | --- |
|  |  | Log reduction | % inactivation | Log reduction | % inactivation | Log reduction | % inactivation | Log reduction | % inactivation | Log reduction | % inactivation | Log reduction | % inactivation |
| 10 µM MB (w/ 30 min light) | RH | 4.4 | > 99.9% | 5.5 | > 99.9% | 2.2 | > 99.4% | 4.3 | > 99.9% | 3.2 | 99.6% | 4.1 | 99.8% |
| 10 µM MB (w/ 30 min light) | RM | 3.5 | > 99.9% | 2.7 | 99.8% | 2.8 | > 99.8% | 4.0 | > 99.9% | 3.4 | 99.9% | 3.2 | 99.9% |
| 10 µM MB (w/ 30 min light) | *R3 | 5.1 | > 99.9% | 5.0 | > 99.9% | 3.3 | > 99.9% | 3.1 | > 99.9% | 3.2 | > 99.9% | 4.1 | > 99.9% |
| 10 µM MB (w/ 30 min light) | *FW | 5.8 | > 99.9% | 5.5 | > 99.9% | 3.6 | > 99.9% | 3.1 | > 99.9% | 3.4 | > 99.9% | 4.5 | > 99.9% |
| 10 µM MB (w/ 30 min light) | FH | 3.8 | > 99.9% | 5.1 | > 99.9% | 3.1 | > 99.9% | 4.2 | > 99.9% | 3.6 | > 99.9% | 4.0 | > 99.9% |
| 10 µM MB (w/ 30 min light) | CM | 3.2 | > 99.9% | 0.4 | 60.5% | 1.3 | > 94.6% | 2.7 | > 99.9% | 2.0 | 97.3% | 1.9 | 88.7% |
| 75°C DH (60 min) | RH | 1.6 | 97.5% | 5.4 | > 99.9% | 2.4 | > 99.6% | 2.9 | 99.8% | 2.6 | 99.7% | 3.1 | 99.2% |
| 75°C DH (60 min) | RM | 1.4 | 95.8% | 4.7 | > 99.9% | 1.1 | 91.8% | 3.4 | 99.8% | 2.2 | 95.8% | 2.6 | 96.8% |
| 75°C DH (60 min) | *R3 | 1.8 | 98.6% | 5.3 | > 99.9% | 2.3 | 99.5% | 3.7 | > 99.9% | 3.0 | 99.7% | 3.3 | 99.5% |
| 75°C DH (60 min) | *FW | 2.1 | 99.2% | 5.5 | > 99.9% | 2.0 | > 99.1% | 3.9 | > 99.9% | 3.0 | 99.5% | 3.4 | 99.5% |
| 75°C DH (60 min) | FH | 1.7 | 98.0% | 5.1 | > 99.9% | 1.0 | > 90.6% | 3.0 | 99.6% | 2.0 | 95.1% | 2.7 | 97.0% |
| 75°C DH (60 min) | CM | 3.1 | 99.9% | 5.0 | > 99.9% | 0.9 | > 88.8% | 2.0 | > 99.9% | 1.5 | 94.3% | 2.8 | 97.1% |

Notes:
<sup>1</sup> Data extracted from inactivation curve (Except for PRCV, SARS-CoV-2 (Lab 2), and MHV 10 µM MB)
<sup>2</sup> Light source for testing MB on PRCV = 12500 lux red light; Light source for testing MB on all other virus types = 50000 lux white light
<sup>3</sup> Due to the limit of detection of the assays used to titer the virus, the highest % inactivation is indicated as >99.9%
<sup>4</sup> RH = Halyard duckbill respirator (Fluidshield-46727), RM = 3M half-sphere respirator (1860), R3 = 3M Panel respirator (1870+), FW = Type II EN 14683 generic face mask, FH = Type IIR ASTM F2100 Level 2 Halyard face mask, CM = Community mask

In addition, we tested the effect of MBL or DH inactivation on MM and FFR straps inoculated with PRCV. MB and DH treatment of FH, FW, and R3 elastic straps resulted in inactivation of nearly 4 logs of PRCV. In contrast, MBL treatment of RH, as well as both MBL and DH treatment of RM respirator straps resulted in infectious titer reductions of less than 3 logs [Supplemental FIGURE S4].

### Community Mask

Cloth community masks (CM) are globally in wide use; therefore, we investigated the decontamination efficacy of MBL and DH on a homemade CM made of a cotton inner and outer layers surrounding two layers of spunbond polypropylene.

Overall, virus recovery from the CM mask (made of cotton inner and outer layers surrounding two layers of spunbond polypropylene) was less efficient than from the FFRs and MMs, possibly due to absorption by the outer cotton layer. This was especially prominent for SARS-CoV-2-inoculated CMs. MBL treatment of CMs resulted in complete inactivation of SARS-CoV-2 and MHV. In contrast, PRCV titers were only reduced by 0.4 log, whereas >5 logs of PRCV were inactivated after DH treatment [FIGURE 5A-D].

**FIGURE 5.**
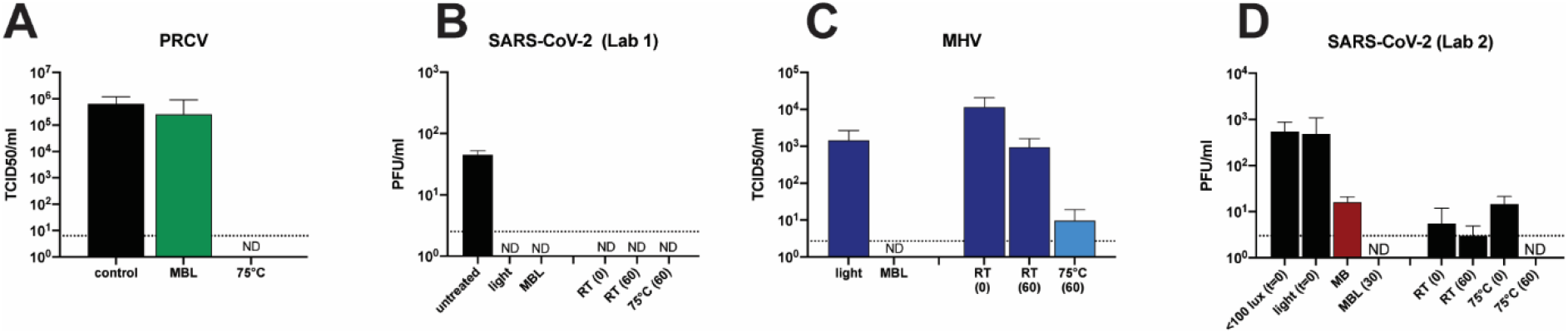
Effect of MBL or DH treatment on MHV, PRCV and SARS-CoV-2 titers inoculated on CMs. Coupons cut from CM were inoculated with 100 ul of PRCV or 10 ul of SARS-CoV-2 or MHV, dried for 20-30 min, and treated with 10 uM MB before exposure to 50,000 lux of light or 12,500 lux of red light (PRCV) for 30 min or exposure to 75°C of DH for 60 min. CM = community mask. ND = not detected. Dotted line indicates limit of detection.

The overall percent reduction in virus titer after treatment across all FFRs/MMs and viruses ranged from 99.8-100% for MBL, and 96.9–99.6% for DH. For the CM this was 88.8% for MBL and 97.2% for DH [TABLE 1].

### Evaluation of Potential Applications of MBL in a Clinical Setting

Three potential applications of MB in a clinical environment were examined. First, since some clinical settings may not have access to bright light, we investigated whether 10 µM MB combined with ambient light would be sufficient to inactivate SARS-CoV-2. Nearly 5-log of SARS-CoV-2 was inactivated by treatment with 10 µM MB and exposure to 700 lux (ambient light generated by light in a biosafety hood) for 60 minutes, and nearly 3-log of virus was inactivated by 10 µM MB in <100 lux of light (biosafety hood with the lights off) [FIGURE 6A]. Second, the possibility of pre-treating respirators/masks with MB was investigated. Coupons were treated with 10 µM MB, dried, and inoculated with SARS-CoV-2 on either the hydrophobic outer layer or the hydrophilic inner layer before exposure to 50,000 lux of light for 30 min. No infectious virus could be recovered from either side of the light-exposed coupons, signifying inactivation of >4 logs of virus [FIGURE 6B]. Intact RM respirators and FH masks were sprayed with 10 µM MB, dried overnight, inoculated with MHV, and exposed to 50,000 lux of light for 30 min. No detectable virus was recovered [FIGURE 6D].

**FIGURE 6.**
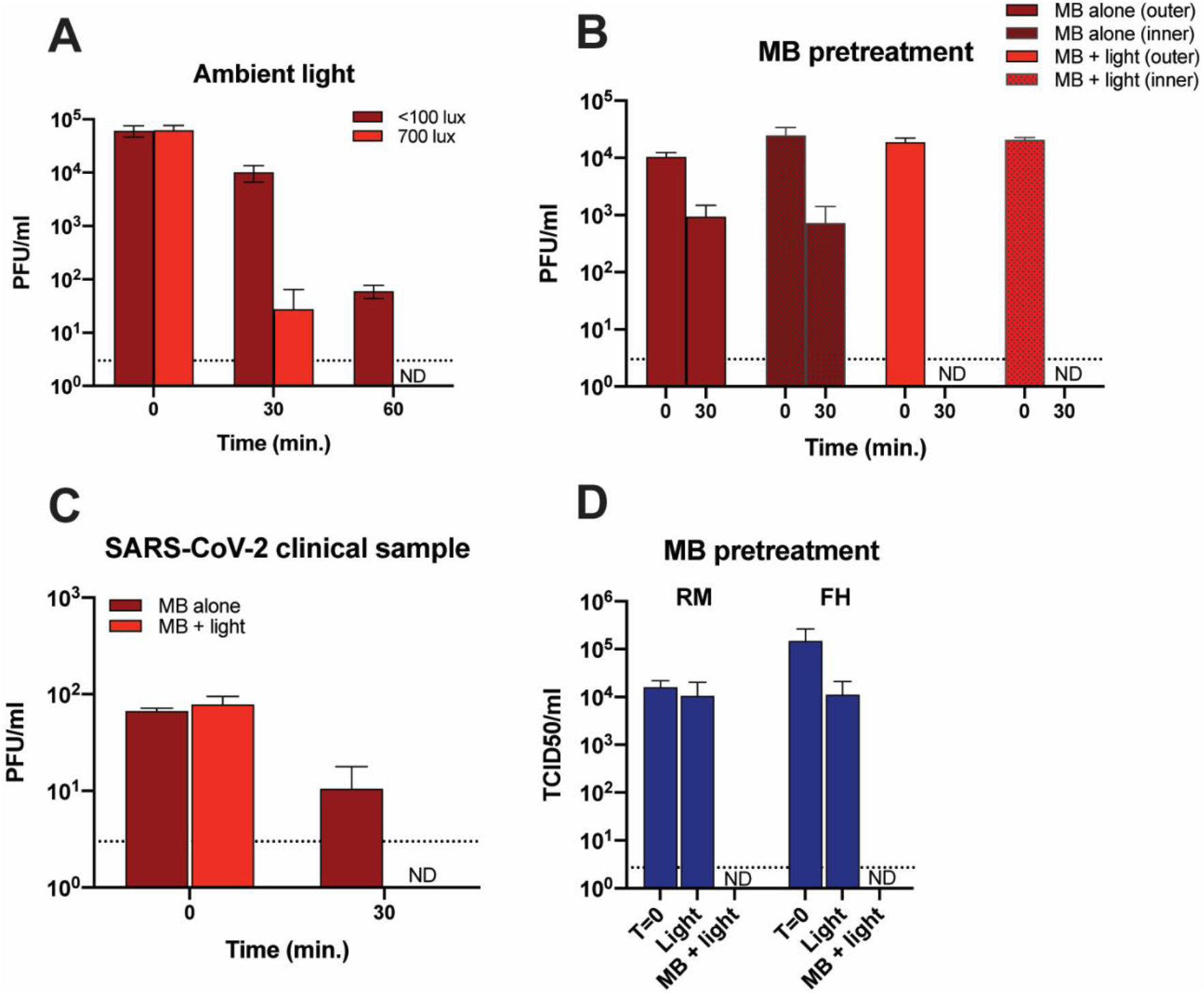
Potential applications of MBL in a clinical setting. **(A)** Effect of low light levels on SARS-CoV-2 inactivation using MB. A 10 µl aliquot of SARS-CoV-2 was applied to R3 coupons and dried for 20 min. Ten microliters of 10 µM MB was added to each coupon before treatment with 700 lux or <100 lux of light. Seven hundred lux is the light level produced by the biosafety hood lights and is used to represent ambient light. **(B)** Effect of MB pre-treatment on SARS-CoV-2 inactivation. Coupons were cut from R3 masks and soaked for 1 hr in 10 µM MB. The coupons were then air-dried in the dark for 2 days before adding 10 µl of virus to either the inner or outer layers. The samples were exposed to light (50,000 lux) for 30 min, and the virus titer determined by plaque assay using 6 replicates. **(C)** Inactivation of a SARS-CoV-2 clinical specimen by MBL. A saliva specimen was obtained from a COVID-19 patient with a titer of 1.1 × 10^5^ PFU/ml for SARS-CoV-2. Ten µl aliquots were applied to coupons cut from a R3 mask and treated with 10 µM MB and exposed to 50,000 lux of light for 30 min. Each virus titer was measured using 6 replicate samples. **(D)** Effect of MB pre-treatment on MHV inactivation using intact masks. Intact R3 and FW masks were pretreated with 10 µM MB by spraying the front and back with a total of 7-8 ml of MB and allowed to dry overnight in the dark. The dried masks were inoculated with MHV and exposed to 50,000 lux of light for 30 minutes, and the inoculated areas were excised before elution and titration. ND = Not detected. FH= Type IIR Halyard face mask. RM= 3M 1860 half-sphere respirator. Dotted line indicates limit of detection.

Lastly, 10 μ l of a sample from a clinical specimen (saliva) with a titer of 1.15 × 10^5^ PFU/ml obtained from a COVID-19 patient was inoculated onto coupons from an R3 respirator and treated with MBL (30 min). No infectious virus was detectable after this treatment indicating that this inactivation method may be effective in a clinical setting where infectious virus may be found in proteinaceous matrixes which would potentially facilitate viability [FIGURE 6C].

### N95 Respirator, Medical Mask, and Community Mask Integrity Testing

Integrity of the FFRs/MMs/CM was assessed using a number of standard test methods to examine if MBL or DH decontamination treatments affected mask integrity. Filtration efficiency, breathability, fluid resistance and fit testing of FFRs/MMs/CM were assessed before and after 5CD. Results were compared to the FDA-authorized VHP+O_3_ decontamination method. The following sections describe each of the integrity test methods and results (see Supplemental Tables S2A and S2B for all integrity test results). Certain test methods are employed for respirator approval/certification by the National Institute for Occupational Safety and Health (NIOSH) and European Notifying Bodies. Other tests, such as for medical masks, are required by the FDA for clearance or for compliance to the European Medical Device Directive.

First, we determined the filtration efficiency of the tested masks/respirators. Filtration efficiency testing measures the percentage of particles of a certain size that are stopped and retained by the mask material. Filtration efficiency testing using sub-micron (count median diameter of 0.075 ± 0.020 µm) sodium chloride (NaCl) particles is typically used to evaluate tight-fitting respirators, while medical masks are typically tested using larger 3-micron droplets containing culturable bacteria (*Staphylococcus aureus*). Sub-micron filtration efficiency testing is the more stringent of the two filtration tests since this method uses smaller and charge-neutralized particles, and a high air flow rate (simulating inhalation at heavy workload). Therefore, the standardized sub-micron testing was used to compare filtration efficiency between FFRs and MMs after decontamination treatments. Bacterial filtration efficiency was assessed only for MMs and CM.

#### Sub-micron Particulate Filtration Efficiency

FIGURE 7A depicts the filtration efficiency of all FFR and MM models before (untreated) and after 5CD. High filtration efficiencies were expected for the FFRs as they were all NIOSH-approved N95 FFRs, which requires ≥95% sub-micron filtration efficiency. All three FFR models surpassed the minimum 95% NaCl sub-micron filtration efficiency requirement before and after 5CD. Untreated FW and FH masks achieved 76% and 86% filtration efficiency respectively, and in general filtration efficiency increased after MBL, DH, and VHP+O_3_ treatments for these masks. Since the CMs were made using materials found in the community, higher variability was observed between individual masks. As seen in FIGURE 7A, CMs achieved 34%, 32%, and 47% sub-micron filtration efficiencies at baseline (before treatment) and after MBL or DH treatments, respectively. Since VHP+O_3_ is not an effective decontamination method for materials containing cellulose, this treatment was not performed on CMs. Overall, MBL, DH, and VHP+O_3_ treatment of FFRs/MMs/CM did not cause any significant differences in the NaCl sub-micron filtration efficiency of the studied models (p>0.01).

**FIGURE 7.**
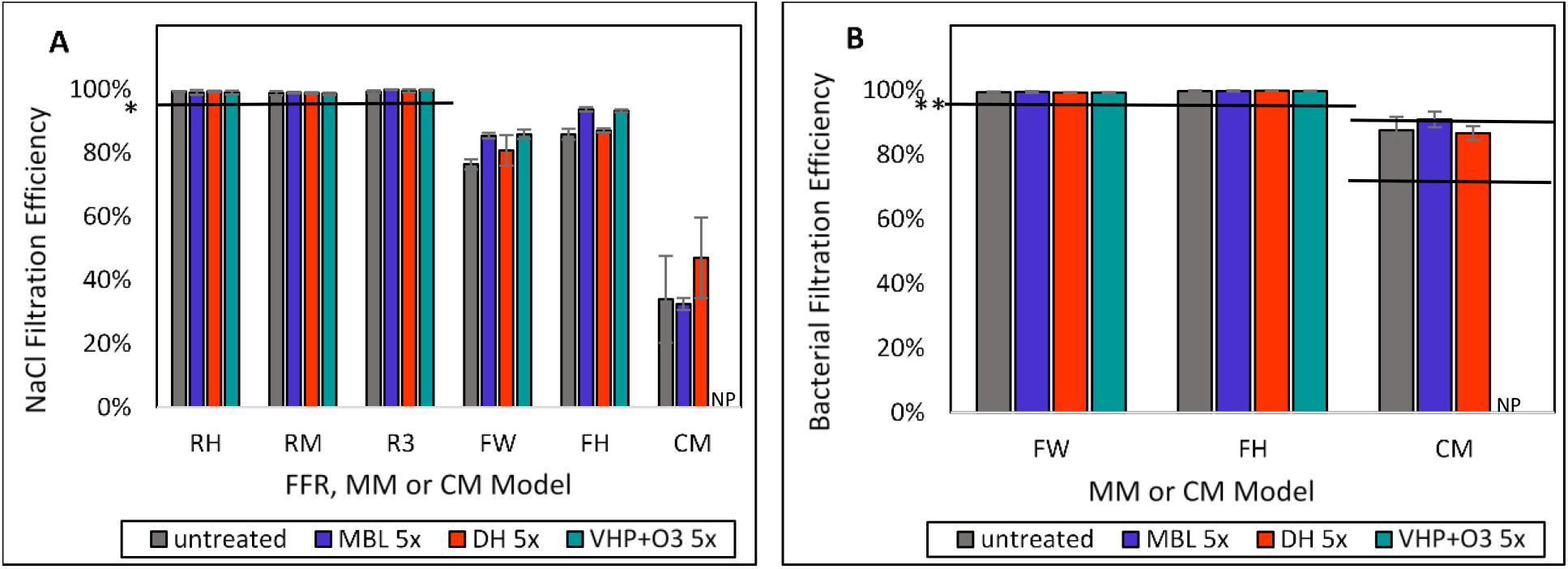
Effect of MBL, DH, and VHP+O_3_ treatments on the filtration efficiency of FFRs/MMs/CM. **(A)** NaCl sub-micron filtration efficiencies of FFRs/MMs/CM before and after 5CD. NaCl sub-micron filtration efficiency is a measure of the ability of a respirator or medical mask to capture aerosolized particles smaller than one micron, expressed as a percentage of particles that do not pass the material at a given velocity or flow rate. Particle size ranges from 0.022–0.259 µm with a count median diameter of 0.075 ± 0.020 µm and a geometric standard deviation (GSD) of less than 1.86 to give a mass median aerodynamic diameter of 0.3 μm and air flow is 85 L/min (which simulates inhalation at heavy workload). **(B)** Bacterial filtration efficiencies of FFRs/MMs/CM before and after 5CD. Bacterial filtration efficiency is the effectiveness of medical face mask material in preventing the passage of aerosolized bacteria suspended in 3 µm droplets, expressed as the percentage that does not pass the mask material at a 28.3 L/min flow rate (similar to human breathing at a light workload). *Horizontal solid line represents the N95 filtration efficiency requirement of ≥95% particle filtration efficiency according to 42 CFR Part 84. **Horizontal solid lines represent the bacterial filtration efficiency (3 µm droplet size) requirement of ≥98% according to EN 14683 Type II and IIR and ASTM F2100 Level 2 and Level 3 MMs; >70% and >90% levels according to CEN CWA 17553 (solid and liquid particles without any biological challenge) and >70% according to AATCC M14 for CM. RH= Halyard duckbill respirator (Fluidshield-46727). RM= 3M half-sphere respirator (1860). R3= 3M panel respirator (1870+). FH= ASTM F2100 Level 2 Halyard face mask. FW= EN 14683 Type II generic face mask. CM=Community mask. NP= not performed.

#### Bacterial Filtration Efficiency (BFE) using 3-micron Droplets Containing Bacteria

Filtration efficiency of medical/surgical mask material is determined using bacterial filtration efficiency (BFE) testing. BFE should be ≥98% according to EN 14683 for Type II and ASTM F2100 for Level 2 medical masks. Both FW and FH masks achieved greater than 98% BFE before and after 5CD of the three different decontamination methods while CM achieved 88%, 91%, and 87% BFE as baseline and after MBL and DH treatments, respectively [FIGURE 7B]. Overall, no significant differences were observed in the BFE values of any of the tested mask models after 5CD with the three decontamination treatments.

#### Breathability Testing

The breathability, or resistance to airflow via inhalation and exhalation, is an indication of the difficulty in breathing through the respirators or masks. Breathability is an important property for FFRs’ and MMs’ wearer comfort. The breathability of FFRs is assessed by inhalation and exhalation resistance tests. According to 42 CFR Part 84 requirements, exhalation resistance should not exceed 25mm H_2_O and inhalation resistance should not exceed and 35 mm H_2_O. EN 149 also requires that the inhalation resistance of respirators (FFP2) at slightly higher flow rate (95 L/min) should be less than 2.4 mbar, or approximately 24 mmH_2_O. Breathability of MMs is determined by a differential pressure test, which is one of the requirements according to ASTM F2100-19 and EN 14683:2019 medical face mask standards. It is also one of the required test methods by FDA for clearance of certain medical masks, including surgical masks.

#### Inhalation and Exhalation Resistance

FIGURE 8A and 8B show inhalation and exhalation resistances of FFRs/MMs/CM before and after 5CD. Results show that all FFR models achieved inhalation and exhalation resistance far below the breathing resistance limits according to applicable standards and regulations, implying that it would not be harder for the wearer to breath through the masks after 5 decontamination cycles. Although assessments of inhalation and exhalation resistance is not commonly conducted on MM materials, these properties were assessed in this study to understand the impact of decontamination processes on MMs, and to compare FFRs with MMs in terms of breathability. MMs demonstrated similar inhalation resistances, but much lower exhalation resistance values compared to FFRs. Inhalation and exhalation breathing resistances were at least 60% below the allowable thresholds for all FFR models, according to the respective NIOSH 42 CFR Part 84 and EN 149 requirements for breathing resistance of FFRs. Inhalation resistance of all face masks decreased (improved) after 5CD. Notably, inhalation resistance of the CM declined from 8.1 to 4.2 and 3.4 mmH_2_O after MBL and DH treatment, respectively.

**Figure 8.**
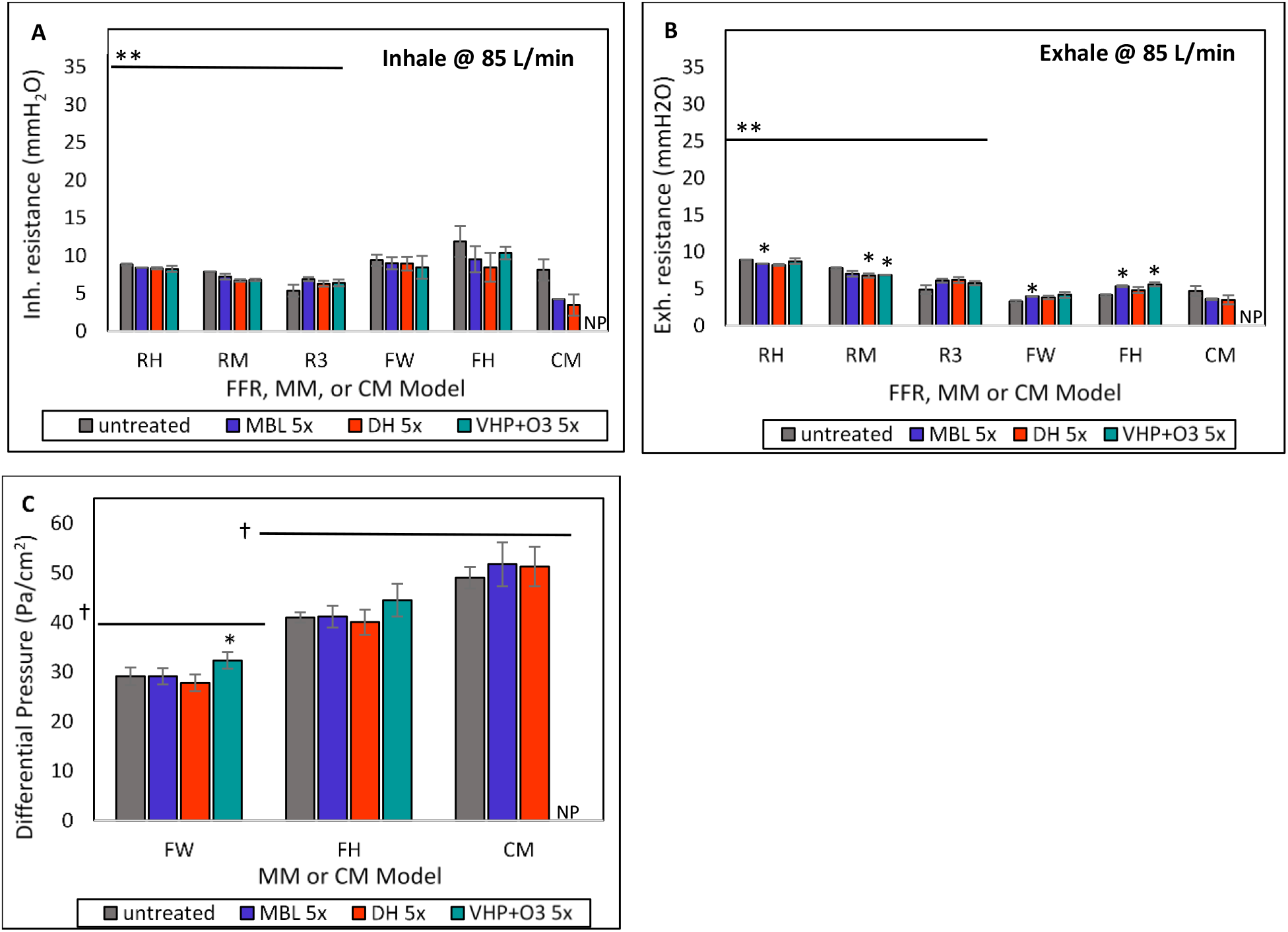
Effect of MBL, DH, and VHP+O_3_ treatments on the breathability of FFRs/MMs/CM. **(A)** Inhalation and **(B)** Exhalation breathing resistances before and after 5CD. The resistance to airflow during inhalation and exhalation is an indication of the difficulty in breathing through the respirators/masks. **(C)** Differential pressure results for MMs/CM before and after 5CD. Pressure drop is a measure of the differential air pressure on either side of the medical mask. *Results from decontaminated FFRs/MMs are significantly different from untreated masks (Student’s t test or Mann-Whitney U test p<0.01). **Horizontal solid lines represent the following breathing resistance standards: Inhalation: ≤35 mmH_2_O; Exhalation: ≤25 mmH_2_O for respirators according to 42 CFR Part 84. Note: EN 149 maximum inhalation resistance at 95 L/min is 2.4 mbar, or approximately 24 mmH_2_O. At higher flow rate according to EN 149, the equivalent breathing resistance may increase slightly but can be similar to the 42 CFR Part 84 maximum inhalation resistance at 85 L/min. Similarly, maximum resistance at 95 L/min is 2.4 mbar for inhalation and 3.0 mbar for exhalation according to CEN CWA 17553 for cloth masks. †Horizontal solid lines represent the maximum allowed differential pressure in the following standards: <40 Pa/cm^2^ according to EN 14683:2019 Annex C for Type II; <58.83 Pa/cm^2^ according to ASTM F2100 for Level 2; and ≤70 Pa/cm^2^ according to CEN CWA 17553 for CMs. RH= Halyard duckbill respirator (Fluidshield-46727). RM= 3M half-sphere respirator (1860). R3= 3M panel respirator (1870+). FH= ASTM F2100 Level 2 Halyard face mask. FW= EN 14683 Type II generic face mask. CM=Community mask. NP: not performed.

#### Differential Pressure

The differential pressure (pressure drop) test measures the differential air pressure on either side of the MM materials using a digital manometer and is a required test method according to ASTM F2100-19 and EN 14683:2019. It is also one of the required test methods by FDA and the European Medical Device Directive for clearance of medical masks. As shown in FIGURE 8C, all of the MMs/CM maintained performance below the maximum allowed differential pressure values before and after 5CD. A slightly higher pressure drop was noted after decontamination with VHP+O_3_ for both MMs Overall, no concerns were reported in terms of breathability after the decontamination treatments. An additional, modified breathability test was performed using a Sheffield dummy that simulated breathing and wear, see Supplemental Figure S5.

#### Fluid Resistance Testing

During a medical procedure, a blood vessel is occasionally punctured resulting in a high velocity stream of blood impacting a medical face mask. Fluid (splash) resistance testing is used to evaluate the resistance of medical face masks to penetration by a small volume (∼2 mL) of a high-velocity stream of synthetic blood. Medical face mask pass/fail determinations are based on visual detection of synthetic blood penetration on the reverse side. Fluid resistance properties of MMs and CM were assessed by both challenging from the inside (inner surface) to investigate their potential source control performance, and from the outside (outer surface) to investigate the protection from splash and sprays that can be encountered during surgeries, patient care, and large droplets (respiratory secretions) that may be generated by other people’s coughs or sneezes.

FW masks demonstrated 60% pass before treatment, and 80%, 50%, and 40% pass when challenged from inside (observed outside) after MBL, DH, and VHP+O_3_ treatments, respectively [FIGURE 9]. Untreated and DH-treated FH masks achieved 90% pass when challenged from the inside. The passing results increased to 100% after MBL or VHP+O_3_ treatments. In addition, untreated, MBL and DH-treated CMs demonstrated 30%, 40% and 50% pass rates respectively, when challenged from inside (observed outside). The fluid resistance of CMs was higher when challenged from the outside compared to the inside. This result was unexpected since the same fabric types were used in the inner and outer surfaces of the CM. However, it may be from the variable and uncontrolled nature of the construction of community masks. Furthermore, it was observed that the decontamination processes increased the fluid resistance of outside materials of both MM models. However, fluid resistance of outside materials of CMs declined after MBL and DH treatments. There were no significant differences between treated masks and untreated masks (Fisher’s exact test).

**FIGURE 9.**
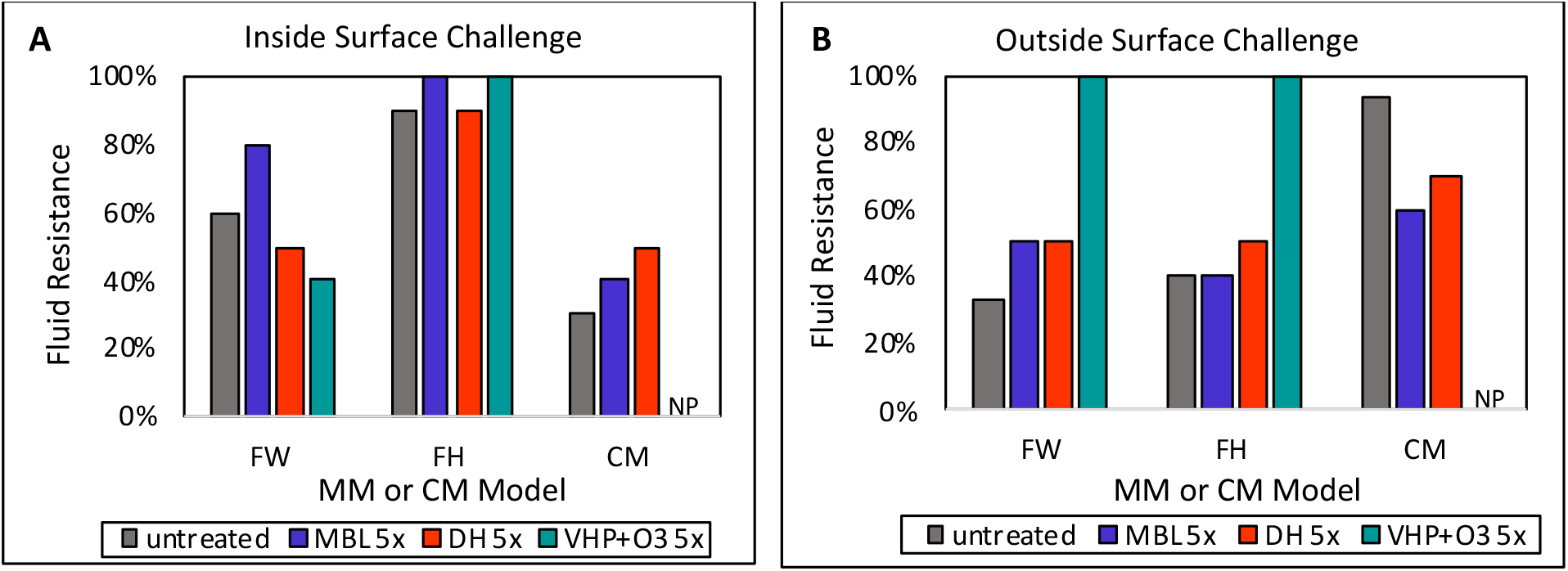
Effect of MBL, DH, and VHP+O_3_ treatments on the fluid (splash) resistance properties of inside and outside surfaces of MMs/CM. Fluid resistance of masks was assessed using a high-velocity (120 mmHg pressure) stream of artificial blood (2 mL). Ten masks were tested per condition, with all masks passing at 100%, meaning fluid penetration was visually undetectable on the opposite side. Masks were either challenged from the inside **(A)** or the outside, with fluid resistance/penetration assessed on the opposite side. FH= ASTM F2100 Level 2 Halyard face mask. FW= EN 14683 Type II generic face mask. CM=Community mask. NP= not performed.

#### Fit Testing

The respirator must fit the user’s face snugly (i.e., create a seal) to minimize the number of particles that bypass the filter through gaps between the user’s skin and the respirator seal. Therefore, fit testing is a critical component to a respiratory protection program whenever workers use tight-fitting respirators, such as N95 FFRs. Fit testing provides a measure of how well a respirator/mask seals around the contours of the face. A good fit ensures exchanged air is filtered through the respirator/mask. Fit was assessed using a PortaCount^®^ PRO+ 8038 machine with human participants (many of whom were healthcare personnel), and their natural anthropometric variations. In addition, advanced headforms that mimicked human head shapes were used to measure fit. The impact of the decontamination method on respirator/mask fit, in terms of fit factors (FF) was compared. Strap/ear loop/tie elastic recovery/tensile strength testing was also performed to investigate discrepancies in fit.

Respirators maintained human participant FF values above 100 after 5 MBL and DH decontamination cycles. However, five cycles of VHP+O_3_ decontamination decreased human fit of RH and RM to the point of failure [FIGURE 10A]. Human fit testing was also performed for MMs and CMs to demonstrate that these types of masks are not designed to ensure a tight fit [FIGURE 10A].

**FIGURE 10.**
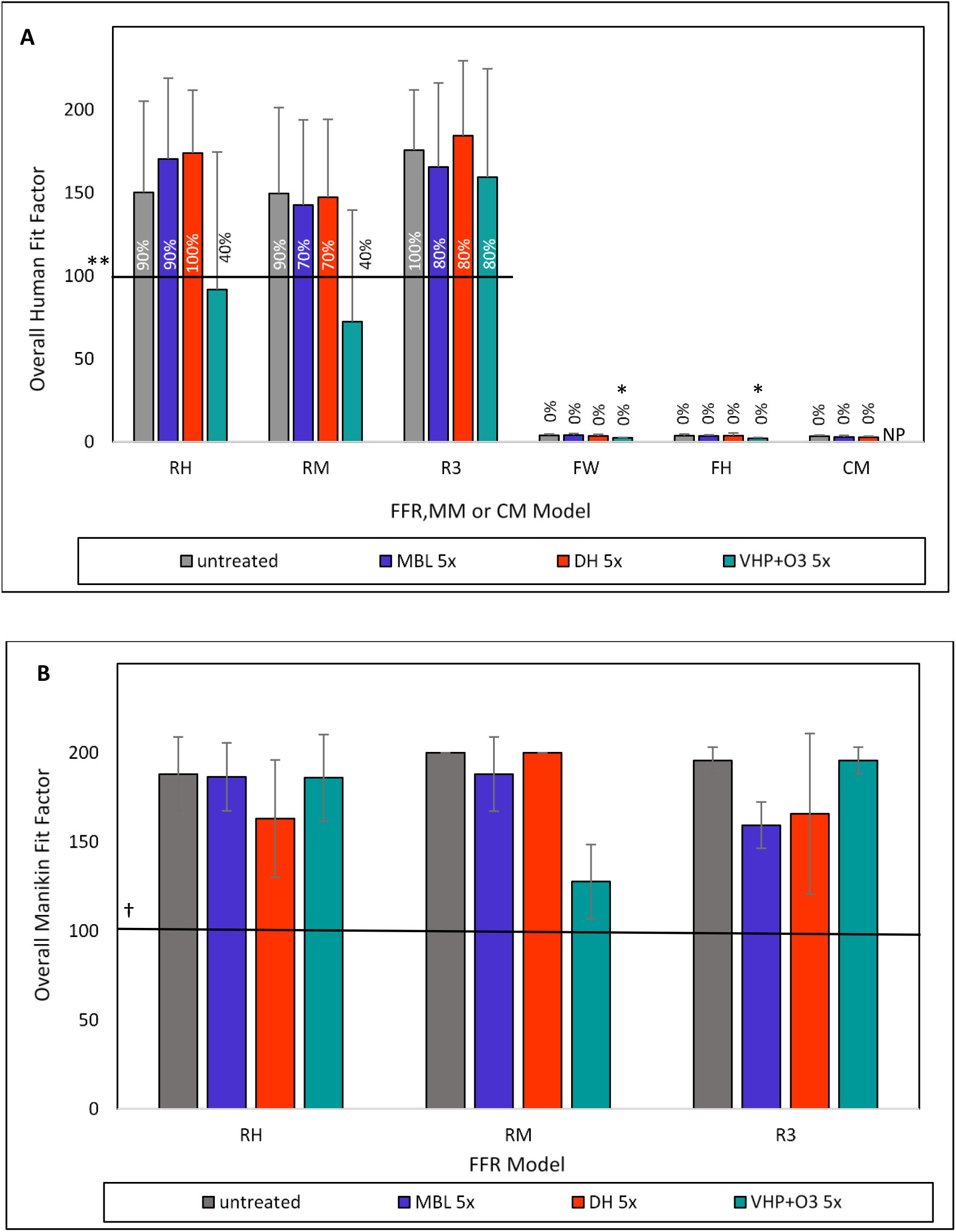
Effect of MBL, DH, and VHP+O_3-_treatments on human and manikin overall fit factor for three FFRs (R3, RH, RM), two MMs (FH, FW) and CM. **(A)** Human fit testing was performed with volunteer participants who adjusted the FFR, MM or CM to achieve the highest fit factors or seal and subsequently performed head movements and remeasured fit or seal. The PortaCount® PRO+ 8038 machine was used to determine overall fit, with human participants and with manikins with advanced head forms. **(B)** Manikin fit factors using advanced, realistic manikin headforms is a reproducible method to test fit without volunteer participants. Its use confirms fit for anticipated facial dimensions. *Indicates significantly different values between treated and untreated FFR, MM or CM at p<0.05, Student’s t-test or Mann-Whitney U test, as appropriate. **Horizontal solid line represents the following standard: ≥100; OSHA 29 CFR 1910.134(f)(10). Percentages on or above each bar represents % of respirators or masks tested that surpassed this standard. While the standard does not apply to face masks, we present the % to note the strong difference between respirator and face mask test results. †Horizontal solid line represents the following standard: Per OSHA 1910.134(f)(10), if the Overall Fit Factor as determined through an OSHA-accepted quantitative fit testing protocol is equal to or greater than 100 for tight-fitting half facepieces, then the fit test has been passed for that respirator. RH= Halyard duckbill respirator (Fluidshield-46727). RM= 3M half-sphere respirator (1860). R3= 3M panel respirator (1870+). FH= ASTM F2100 Level 2 Halyard face mask. FW= EN 14683 Type II generic face mask. CM=Community mask.NP= not performed.

A ‘strong acrid odor’ was noted on some of the VHP+O_3_ treated FFRs and MMs, while ‘not unpleasant slight odor’ was noted on some of the FFR after MBL and DH treatments at one of the fit testing sites. Partial elasticity loss in the VHP+O_3_ treated FFR straps and MMs ear loops were also observed by the wearers, see Supplemental section: Tensile Testing of Elastomeric Straps, Ear Loops, and Ties. Some of the nosepiece foams on VHP+O_3_ treated RMs were discolored (turned from gray to brown). In addition to the discoloration, the nose bridge was more rigid for the three MBL-treated RMs.

Manikin fit factors using an advanced, realistic manikin head, resulted in similar overall passing of the OSHA 1910.134(f)(10) criterion of 100 FF for all three FFRs (11) [FIGURE 10B]. The differences observed between human fit and manikin fit can be attributed to the variability in testing procedures and human facial dimensions and generally the challenge of FFRs to universally fit the wearer.

## DISCUSSION

We observed that methylene blue and light applied to FFR and MM completely inactivates SARS-CoV-2. Importantly, MBL, as a new proposed decontamination method, did not cause any significant changes in any of the filtration, breathability or fluid resistance properties investigated in this study after 5 decontamination cycles, and our results are consistent with filtration efficiency seen after VHP+O_3_ treatment. The additional finding that MB-pretreated FFRs and MMs continue to inactivate coronavirus provides a potential opportunity for healthcare providers and other front-line workers to safely re-use their PPE, as well as supply active viral inactivation during wear. MB is available globally and can be applied using spray bottles. This study demonstrates viral inactivation using MB combined with 50,000 lux of broad-spectrum light (∼1/3 of illumination on a sunny day). Bright light treatment can be supplied by simple white light lamps sold in hardware stores. Furthermore, in a small preliminary experiment, even MB exposed to ambient light (700 lux) and low light (<100 lux) inactivated SARS-CoV-2. This finding needs to be further studied but suggests that MB can work using ambient light available in hospital and clinic settings. We did observe, however, that lower light intensities require a longer light exposure duration for efficacy.

Identifying methods to decontaminate PPE to protect healthcare personnel is paramount. A recent study demonstrated recovery of viable virus from FFR material inoculated with viral loads of SARS-CoV-2 and stored at 20°C with 40-50% relative humidity for 21 days. (12) These common conditions in health care settings demonstrate the importance of having an inexpensive, simple, and robust means of decontamination of PPE like MBL that can retain functionality and performance. In our study, PRCV was inoculated under the outer layer of MMs and FFRs to mimic the worst-case scenario of high-titer virus penetrating the outer layer of a mask. MBL robustly inactivated >5 logs of virus for both MMs and 2 of the FFRs, suggesting that both MB and light were able to sufficiently penetrate the outer layer of the PPE. The exceptions were RM (2.7 log or 99.8% reduction) and CM (0.4 log or 60.5% reduction). RM is a half-sphere with a more rigid shell which may have blocked some of the MB or light from penetrating to the inner layer. Fisher et al. saw a similar finding with UVC light penetration, which was impacted the further into the filter material the light passed. (13)

Consistent with previous studies (14), the DH and VHP+O_3_ treatments did not alter the filtration efficiency of masks and respirators tested in this study. MB and the DH decontamination can now be added to the rare category of decontamination methods that do not violate mask integrity for the models tested (including fit), which thus far only included the gold standard VHP (3,9,14-16-). Although the FDA approved a few VHP systems for PPE decontamination, they are not easily obtainable in low-resource settings. Various studies have also investigated the impact of heat on the filtration efficiency and physical deformation of FFR or filter media and demonstrated that higher temperatures could negatively affect integrity (13,17,18).

Dry heat yielded inconsistent viral inactivation in our study possibly owing to evidence that humidity plays a role in heat inactivation (19). The finding that DH on CM materials yielded better viral inactivation may be due to the ability of the cotton materials to rise to inactivation-level temperatures faster than the synthetic FFR and MM materials, although we did not test the material surface temperature during the heating to prove this. In one study, improved inactivation was demonstrated with moist heat (100% relative humidity) reducing initial SARS-CoV-2 titers almost 5-logs at 75°C for 30 minutes (19). Our rationale for testing 75°C DH without humidity related to testing the simplest method which could be replicated in any location where standard ovens exist without the complexity of regulating or delivering a certain amount of humidity.

We decided to compare a CM in this study to provide first definitive analysis in the same study format used for respirators and medical-grade masks. The tested CM performed surprisingly well in some of the integrity tests. This is likely due to the double cotton layer combined with two layers of polypropylene spunbond fabric. Not all CMs in use around the world contain these multiple layers, especially non-woven layers. The heterogeneity of materials and design, and lack of standardization of CM used for source control make generalized conclusions difficult. When testing MBL virucidal activity on CM, we observed inconsistent results when virus was inoculated under the cloth layer. This could be due to absorption of MB into the cotton layer. Alternatively, CM material color or finishing dye chemicals potentially impeded MB efficacy through light absorption. These hypotheses should be tested using alternative CM fabrics, colors, and models.

Our studies on MBL, DH, and VHP+O_3_ decontamination were constructed to parallel other studies investigating the impact of heat and VHP+O_3_ on the filtration efficiency and physical deformation of FFR or filter media. Previous studies evaluated meltblown polypropylene filter media using NaCl aerosol testing and found that filtration efficiency remained ≥95% after twenty treatment cycles consisting of 75°C for 30 min (14,17,20). On the other hand, Viscusi et al. (15) autoclaved two FFR models at 121°C/15 psi for 15 or 30 minutes and reported that filtration efficiency of both models decreased below minimum NIOSH requirements. A study investigating dry heat at lower temperature settings (110°C) reported that reduction of filtration efficiency was FFR model-specific (16). Previous studies also reported that VHP did not result in the reduction in the filtration efficiency of any of the N95 FFR models tested, while showing significant inactivation of spores, bacteriophages, and SARS-CoV-2 virus (3,9,14,16,20). Battelle CCDS (Critical Care Decontamination System)™ was approved by the FDA as the first emergency use authorization (EUA) for decontamination system for FFRs (3). In Battelle’s system, the 3M 1860 model (RM) was shown to maintain filtration performance for 50 treatment cycles of VHP treatment using the Clarus® R HPV generator (utilizing 30% H_2_O_2_) (3). Additionally, FFR fit was shown to be unaffected for up to twenty VHP treatments cycles using a manikin headform which was assessed on one FFR model without donning and doffing between cycles.Strap degradation, however, did occur after twenty treatment cycles (3). In addition, the FDA has issued an EUA for the SSS VHP N95 Respirator Decontamination System manufactured by Stryker Sustainability Solutions where reprocessing is limited to three cycles (2).

Fischer et al. (14) incorporated test subject quantitative respirator fit testing of intact FFRs using a PortaCount® following each cycle of heat treatment and wear for two hours and reported that the mean fit factor of six fit tests remained >100 for both one and two treatment cycles, but fell slightly below 100 after three dry heat (70°C) cycles. They also reported that one FFR did not have any physical deformation. The same study reported that the mean fit factor of six fit tests remained acceptable (according to OSHA criterion) following three VHP treatment cycles.

We also collected additional measures related to the physical appearance, physical comfort, and user acceptance of decontaminated FFRs/MMs. The purpose of these measures was to determine whether different decontamination methods impact FFRs and MMs in ways beyond physical fit, which could affect perceptions of healthcare personnel in terms of their use of decontaminated devices and ultimately if the use of decontaminated devices would affect HCPs’ performance and delivery of care. We plan to analyze and report upon this component of the study in a future publication.

There were limitations to this study. We only tested 3 types of FFRs, yet there are approximately 500 N95 FFR models approved by NIOSH and many medical masks around the world. Each FFR, MM, or CM model has distinct filter/strap/ear loop/tie materials and design characteristics (e.g., surface area, dead space, nose padding). The differences in materials and design characteristics among all available FFR/MM/CM models result in variable outcomes for decontamination efficacy and effect of decontamination on FFR performance. CDC recommends that a decontamination method’s effectiveness be evaluated for specific FFR models in collaboration with the manufacturer, and if needed, a third-party laboratory (21).

Another limitation within our study was that our primary inactivation studies were performed with direct application of a fixed quantity of MB onto coupons and only two of our experiments tested the spraying of MB onto a whole mask or the soaking of PPE material into a solution of MB. We attempted to normalize the volume of MB applied to the coupons to the amount of per square surface area. For example, if 7-8 mL of MB solution was sprayed onto an intact mask (4 sprays on outer and 2 sprays on inner surface), based on the surface area of a 3M N95 1860 respirator (RM), this would result in 10-30 ul of MB for the each 1 cm^2^ coupon.

In our clinical specimen experiment, we demonstrated SARS-CoV-2 inactivation with MBL. Obtaining whole masks worn by healthcare personnel who cared for COVID-19 patients was not in the scope of our study, yet would add strength to our finding that a saliva specimen from a COVID-19 positive patient was inactivated by the MBL treatment.

All labs performed their studies with ‘biological triplicates’ – three like-masks or respirators from the same respective manufacturing batches were used for every mask tested. We attempted to enhance and strengthen our findings by leveraging four different virology testing sites using three different coronavirus species, including two surrogate coronaviruses. Our findings with the surrogate coronaviruses allow future studies to test diagnostics and therapeutics at lower levels of biocontainment than required for SARS-CoV-2. We believe that conducting our simultaneous experiments across two continents using the same methodology but in different labs, as well as conducting additional experiments using a SARS-CoV-2 clinical sample provides generalizable knowledge. The heterogeneity of light administration methods replicated practical light scenarios. The ovens used in the studies were all different and only standardized for the temperature administered (75°C). The use of alternative heat and alternative light producing methods parallels what we would expect in the real-world settings – no standard products, but ability to standardize a protocol.

The biocompatibility of MB on masks was not tested in this study yet the concentrations of MB – 10 μ M – used to spray onto the masks or used to drop onto mask coupons, were far below the clinically used MB concentrations administered intravenously for methemoglobinemia (1 mg/kg) (5), orally as third line therapy for malaria (22), or applied intranasally for *Staphylococcus aureus* (23). The amount of MB sprayed onto each mask using our 10 µM solution was <0.026 mg. Our team will be testing the amount of MB that a wearer might inhale during the course of an 8-10 hr shift wearing a mask. Although human fit testers of the MBL-treated masks did not experience any negative effects, more rigorous inhalational biocompatibility tests need to be done.

We did not replicate multiple real-life PPE donning and doffing over multiple shifts or extended wear which could affect FFR fit and performance over time. Degesys et al. tested the real time fit using models we used in our study. The 3M 1860 maintained fit for more than 5 donnings (24). In addition, off-gassing of decontamination chemicals was not evaluated although anecdotally, some wearers of the VHP+O_3_ treated masks complained of an “acrid” or “acidic” smell while some wearers of the MBL-treated masked claimed that there was a “mild” and “calming” aroma appreciated. Human factors assessments, including user acceptance of decontaminated FFRs, MMs, and CMs by HCPs are underway. Further testing of the MBL method for user acceptance and workplace compatibility are also being explored.

We demonstrate that MB activated by broad-spectrum white or red light effectively inactivates coronaviruses including the novel SARS-CoV-2 on typical medical mask surfaces without affecting mask material integrity, or alterating mask fit and filtration characteristics. This series of studies provides evidence that MB can be applied to the surface of a FFR or MM and illuminated by a light source as a disinfection technique, enabling safer reuse of single-use disposable respirators and masks in shortage situations. One could envision using this decontamination process after each work shift in a healthcare setting. Integrity was maintained for the FFR models tested in this study after 5CD, however, the variability in the the design and materials (e.g., straps and earloops) may still lead to mask performance changes with successive donnings and doffings that will still need to be assessed to ensure adequate fit of the respirators. Residual MB remaining on the mask surface activated by ambient white light could also provide a novel means of continual disinfection of viral particles which become adherent to the mask surface in-use. Indeed, under ambient lighting condition, the surface of the masks treated with MB could produce up to 3.2×10^15^ molecules of singlet oxygen per second (Supplemental Table S6). Other potential uses include application of MB to face coverings used in low resource medical and general community settings, in order to enhance protection, not only of persons in proximity to the face covering user, but also to increase protection of the user from airborne virus. In the medical setting, on site, MBL spot treatments of PPE touch points prior to doffing may also decrease accidental contamination and may potentially provide increased protection of healthcare and laboratory workers. A further series of experiments intended to support the implementation of these additional uses of MB and light are underway.

## CONCLUSION

We provide the first evidence that methylene blue and light can inactivate human coronavirus on FFRs and MMs commonly worn by healthcare personnel and essential workers without decreasing performance and fit. Our findings provide a recipe for easily accessible, inexpensive, effective PPE re-use and afford an opportunity for utilization in high- and low-resourced settings to address issues of global supply shortages and reduced time of decontamination relative to VHP+0_3_. Residual MB remaining on the mask surface activated by ambient white light could also provide a novel means of continual disinfection of viral particles which become adherent to the mask surface in-use, continually reducing wearer exposure to SARS-CoV-2. We are investigating applying MBL to a wider range of high-threat pathogens.

## MATERIALS AND METHODS

### Respirators and Masks

In this study, three FFRs - Halyard Fluidshield-46727 duckbill respirator (RH, NIOSH Approval Number: TC-84A-7521) (O & M Halyard,), 3M 1860 half-sphere respirator (RM, NIOSH Approval Number: TC-84A-0006), and 3M 1870+ panel respirator (R3, NIOSH Approval Number: TC-84A-5726) (3M), and three MM models - generic EN 14683 Type II medical mask (FW), Halyard ASTM F2100 Level 2 procedure mask (FH) (O & M Halyard), and a homemade community cloth mask (CM) - were tested [TABLE 2.]. All of the FFRs used in this study are surgical FFRs. Surgical FFRs are NIOSH-approved particulate respirators that have also been cleared by the FDA as medical devices. The CM consisted of an inner and outer cotton layer surrounding two layers of polypropylene spun-bond fabrics. See Supplemental Table S1 for details on which labs and testing sites respirators and masks were sent to for decontamination and testing.

**TABLE 2.**
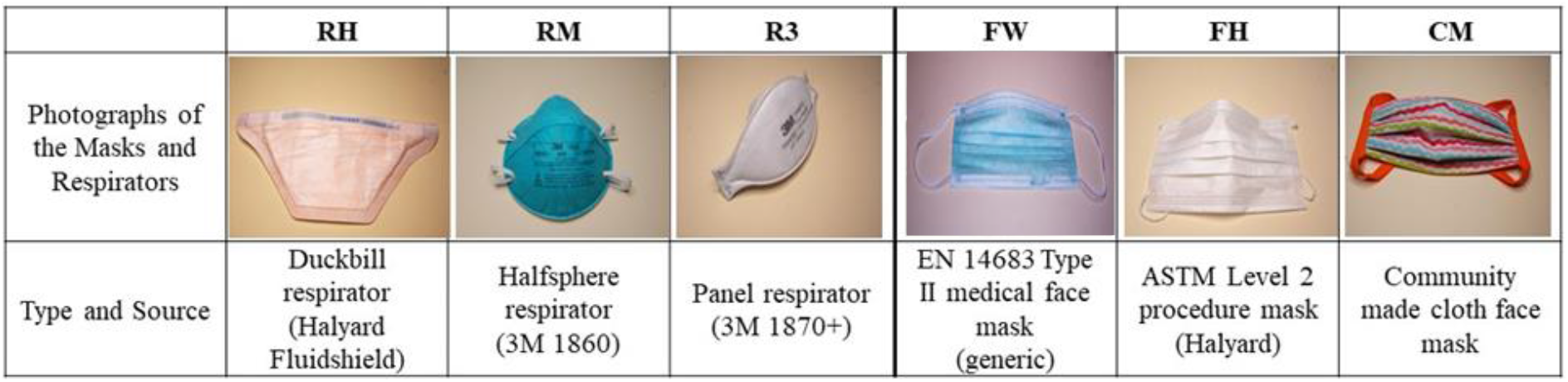
Masks and Respirators used in this study.

|  | RH | RM | R3 | FW | FH | CM |
| --- | --- | --- | --- | --- | --- | --- |
| Photographs of the Masks and Respirators |  |  |  |  |  |  |
| Type and Source | Duckbill respirator (Halyard Fluidshield) | Halfsphere respirator (3M 1860) | Panel respirator (3M 1870+) | EN 14683 Type II medical face mask (generic) | ASTM Level 2 procedure mask (Halyard) | Community made cloth face mask |

### Viruses

SARS-CoV-2 was obtained from Dr. Darryl Falzarano (VIDO) (GISAID accession ID: EPI_ISL_425177) used in Lab 2 or from a patient at the George Washington University Hospital used in Lab 1 and was propagated in Vero CCL-81 cells. Viral titers were determined by plaque assay and typically reached 1.0-6.6 ×10^6^ PFU/mL. A SARS-CoV-2 clinical saliva specimen with a titer of 1.1 × 10^5^ PFU/ml was obtained with the University of Calgary Conjoint Health Research Ethics Board approval (ID# REB20-0444). The recombinant murine hepatitis virus (MHV) stock (rA59-E-FL-M) was described previously (25). Porcine respiratory coronavirus (PRCV), a spike gene deletion mutant of transmissible gastroenteritis virus (TGEV) and a member of the *Alphacoronavirus 1* species (26,27), was used as a SARS-CoV-2 surrogate. PRCV strain 91V44 (28) was passaged three times on ST cells. Both MHV and PRCV were used as SARS-CoV-2 surrogates.

### Tissue Culture

Murine Delayed Brain Tumor (DBT) cells were cultured in MEM supplemented with 1% GlutaMAX, 1% HEPES, 1% NEAA, 10% Tryptose Phosphate Broth and 10% Fetal Bovine Serum (FBS). Vero CCL-81 cells were purchased from ATCC and cultured in DMEM supplemented with 10% FBS, 1% GlutaMAX, 1% Pen/Strep, and 0.1% Amphotericin B. Alternatively, Vero CCL-81 cells were cultured in MEM supplemented with 2 mM L-glutamine, 1 mM Sodium Pyruvate, 1x NEAA, 1% Pen/Strep, 0.1% Amphotericin B, and 10% FBS. Vero E6 cells (ATCC) were cultured in DMEM supplemented with 10% FBS and 1% GlutaMAX. Swine testicular (ST) cells were maintained in MEM supplemented with 5% FBS, 1% sodium pyruvate, and antibiotics (100U/ml penicillin, 0.1 mg/ml streptomycin, 0.05 mg/ml gentamycin). All cells were grown at 37°C in a humidified incubator with 5% CO2.

### Light Sources for Methylene Blue Testing

Light boxes developed at Colorado State University were used at Seattle Children’s Research Institute, George Washington University, University of Calgary and Nelson Laboratories containing the 4000K Husky LED lights (model# K40187). Husky LEDs were also used at the University of Alberta but were 3500K. The University of Liège and Centexbel used a light box of their own design containing horticultural lamps for their studies. Luminescence was verified using light meters in all laboratories (Latnex, model LM-50KL; DeltaOHM, Model HD2102.2; or Cooke, Model CK-CL400; see Supplemental section).

### Virus Inoculation and Elution

Masks were cut into 7-10 mm^2^ coupons and inoculated with the maximum available titer of SARS-CoV-2 or the surrogate coronaviruses (MHV or PRCV). The volume of the inoculum for SARS-CoV-2 and MHV was 10 μ L. Virus was added to the outer layer of each coupon (or inner layer where specified) with a pipette and dried for 20 mins before inactivation treatments were initiated. For elution, coupons were soaked in media in a 1.5 ml microtube (MHV, SARS-CoV-2 experiments) or 15-ml tube (PRCV experiments). Tubes were vortexed or rocked on an orbital rocker for 10-20 minutes. Alternatively, 100 μ l PRCV was injected under the respirators/masks outer layer using an ultra-fine insulin needle. After decontamination, 34 mm^2^ coupons were cut from the masks, placed in a 15-ml tube containing media and vortexed. Remaining infectious virus was quantified by median tissue culture infectious dose (TCID_50_)or plaque assays.

### Methylene Blue Treatment

MB was obtained from Sigma (M9140), ThermoFisher Scientific (J60823) or American Regent (0517-0374-05). Stock solutions were prepared using ultrapure distilled water. After inoculation of the virus and a 20 min drying step, 10-30 μ l of MB at the indicated concentrations was added to the coupons and exposed to 50,000 lux of broad-spectrum light or 12,500 lux of red light for varying time points. Broad-spectrum light and red-light sources were based on the local labs’ method for administering adequate light energy for MB activation. And 12,500lux of red light roughly equates to the same amount of light energy within the MB excitation wavelength that 50,000 lux of white light yields. Intact inoculated masks were sprayed with 7-8 ml of 10 μ M MB and allowed to dry for 30 minutes protected from light before exposure to 12,500 lux of red light for 30 min. For dark control, coupons and masks were left in biosafety hood with the light off or covered by aluminum foil (<100 lux).

For the pre-treatment tests, R3 coupons were soaked with 10 µM MB for >1 hour and dried 2 days protected from light. SARS-CoV-2 was spotted on outer or inner mask layers, dried for 20 minutes before exposure to 50,000 lux of light for 30 min. Virus was eluted from the coupons and quantified by plaque assay. Intact RM and FW received 6 sprays on the front and 2 in the back (8 ml total) and dried overnight. MHV was added to three points on the outer surface, dried for 20 minutes and exposed to 50,000 lux of light for 30 min. The inoculated areas were excised from the mask, eluted, and quantified by TCID_50_ assay.

### Dry Heat Treatment

Inoculated coupons were placed in a tissue culture plate and incubated at the indicated temperatures in a dry heat oven (IsoTemp, Fisher Scientific, VWR Dry-Line 56-Prime oven or Lab-line L-C oven with a DataChart 1250 temperature probe). A calibrated analog thermometer was used to verify temperature if a temperature probe was not available. Intact inoculated FFRs and MMs were placed horizontally on a metal frame in a dry oven (M-Steryl, AMB Ecosteryl Company) containing two calibrated Pt100 sensors to record the temperature throughout the experiment to ensure correct exposure conditions.

### PPE Integrity Testing

Samples were tested as-received (‘untreated’), with DH, with VHP+O_3_ and with MBL. All decontaminated samples were treated off-site (Nelson, 4CAir, University of Liege) before reaching the integrity labs for testing.

### Filtration Efficiency Testing

Filtration was assessed using NaCl sub-micron particles for FFRs, MMs, and CM; and using 3-micron droplets that contain bacterial challenge for MMs and CM. Although medical face masks are not typically tested using NaCl sub-micron particles, performing the tests with this challenging-size particle allowed comparison of filtration performance between masks and respirators. Sub-micron particulate filtration efficiency is a measure of the ability of a respirator or medical mask to capture aerosolized particles smaller than one micron, expressed as a percentage of a known number of particles that do not pass the material at a given face velocity for flat samples or flowrate for whole article testing. Untreated and decontaminated respirators were tested for filtration efficiency using a modified version of the NIOSH Standard Test Procedure (STP) TEB-APR-STP-0059 (29), using an Automated Filter Tester (CERTITEST®, Model 8130, TSI, Inc.) and according to the National Personal Protective Technology Laboratory (NPPTL) Decontaminated Respirator Assessment Plan (11). In sub-micron testing, charged-neutralized particle size ranges from 0.022–0.259 µm with a count median diameter of 0.075 ± 0.020 µm and a geometric standard deviation (GSD) of less than 1.86 to give a mass median aerodynamic diameter of 0.3 μm, and air flow is 85 L/min (which simulates inhalation at heavy workload). Respirator testing was completed at the NIOSH NPPTL. Results were evaluated against NIOSH performance criteria according to 42 CFR Part 84 for FFR approvals (minimum 95% filtration efficiency) (30). Untreated and decontaminated medical face masks and community masks were also tested following the same method at Nelson Laboratories.

The bacterial filtration efficiency (BFE) test is a conventional method to measure filtration efficiency of medical face masks (31). BFE was measured using the bacteria *Staphylococcus aureus* (diameter: 1 μm) as the challenge organism. A suspension of *S. aureus* was aerosolized using a nebulizer to give a challenge level of 1700–3000 colony-forming units (CFU) per test as specified by the ASTM F2101 method (32). The bacterial aerosol is a water droplet containing the bacteria and not an individual bacterial particle. The particles were not charge neutralized for testing. The aerosol sample was drawn through a test sample clamped into the top of a 6-stage Andersen sampler with agar plates for collection of the bacteria particles at a flow rate of 28.3 L/min for 1 min. The design of 6-stage Andersen sampler is based on the human respiratory tract, where all airborne particles greater than 0.65 μm are classified aerodynamically. The flow rate of 28.3 L/min is similar to human breathing flow rate (at light workload) to obtain deposition of particles in different stages of the Andersen sampler. Aerosol droplets generated in this test ranges from 0.65 to 7 µm with a mean particle size of approximately 3.0 µm. As previously mentioned, BFE was employed on masks only. It was performed on untreated and decontaminated MMs and CMs at Nelson Laboratories and Centexbel. Results were evaluated against performance criteria according to EN 148683 Type II and ASTM F2100 and Level 2 medical masks (≥98% 3 µm bacterial droplet filtration efficiency), CEN CWA 17553 (≥70% and ≥90% 3-micron solid or liquid particles without biological challenge) (33) and AATCC M14 (20) (>70% 3-micron latex spheres) for CM.

The filtration efficiency of fibrous filter materials is controlled by factors including, aerosol charge, particle size distribution, face velocity and filter material charge. When NIOSH sub-micron and BFE test methods are compared, NIOSH approval testing is considered as more stringent or worst-case method, because of the use of charge neutralized aerosol size close to the most penetrating particle size at relatively higher flow rate (face velocity), to produce maximum penetration or conservative filtration efficiency.

### Breathability Testing

Breathability was assessed using inhalation and exhalation breathing resistance measurements according to the NIOSH standard testing procedures according to 42CFR Part 84 for the all of the respirators and masks (30, 34) and pressure drop measurements for masks only (ASTM F2100 and EN 14683). An additional breathability assessment using clause 7.16 of the EN 149 standard for European respirators was used and is termed “Sheffield Dummy Air Flow Differences” or Sheffield AFD (EN 149).

Inhalation and exhalation resistance of devices were tested using NIOSH STPs (TEB-APR-STP-0007 and TEB-APR-STP-0003). The results in mmH_2_O were recorded and evaluated against NIOSH performance maximum limits for FFR approvals (25 mmH_2_O for exhalation and 35 mmH_2_O for inhalation) at approximately 85 ± 2 L/min airflow. Respirator testing was completed at the NIOSH NPPTL while MMs and CMs were tested at the Nelson Laboratories.

The differential pressure (*delta p*) testing of untreated and decontaminated samples was performed according to ASTM F2100 and EN 14683 and measured the differential air pressure on either side of the mask using a digital manometer at Nelson Laboratories and Centexbel Laboratories. The *delta p* values were reported in mm water/cm^2^ (required units for ASTM F2100) and Pa/cm^2^ (required units for EN 14683) of test area and evaluated against ASTM F2100 Level 2 and EN 14683 Type II requirements for and masks (ASTM F2100 <6.0 mm H_2_O/cm^2^ and <40Pa/cm^2^ for Type II).

### Fluid Resistance Testing

Testing the fluid penetration resistance (resistance to splash and spray) by synthetic blood is one of the requirements of FDA surgical mask clearance in the U.S. for all three different levels of masks defined in ASTM F2100. It is also required by EN 14683 for Type IIR masks only. The synthetic blood penetration testing of untreated and decontaminated samples was performed on medical face masks and community masks at Nelson Laboratories and Centexbel according to ASTM F1862 or ISO 22609:2004 at 120 mmHg pressure which is required for ASTM F2100 Level 2 masks and EN 14683 Type IIR masks. For each mask type, five samples were tested with the outside facing the synthetic blood to determine the barrier resistance and five samples were tested with the inside facing the synthetic blood to assess the source control properties. The percentage of passing samples (no visible penetration) was calculated and compared to ASTM F2100 requirements (29 out of the 32 passing results).

### Fit Testing

#### Human Fit

Fit testing was conducted with the PortaCount® Pro+ 8038 (TSI Inc., Shoreview, MN, USA) at two study sites: Stanford University, and the University of Calgary. The human fit testing was determined to be exempt from ethics board review by both the Research Compliance Office, Stanford University (March 23, 2020) and the Conjoint Health Research Ethic Board, University of Calgary (June 12, 2020). All fit testing of masks was done with OSHA-approved protocols for testing N95. However, sample sizes and study designs differed between sites. At Stanford University, fit testing of test masks was performed on a human model (LC) using the OSHA test protocol of the PortaCount Pro+ 8038. At the University of Calgary, three groups of five healthcare personnel completed fit tests a repeated-measures design, with respirator type and between-subjects factor. At both sites, the fit test protocol consisted of eight dynamic tasks: normal breathing, heavy breathing, turning head side-to-side, head up-and-down, talking, grimace, bend over and normal breathing. Each cycle of tests was performed twice for each mask and Fit Factor (FF) was calculated.

In addition to these measures, participants at the University of Calgary (including 15 additional healthcare personnel who participated in a separate study arm focused on surgical/procedural and cloth masks; total N = 30) provided data on measures related to the physical appearance, physical comfort, and trustworthiness and acceptance of the decontaminated masks. Data from these measures, which is not included in the current manuscript, will be presented in a separate publication.

#### Manikin Fit

The static advanced headform fit testing (29) was used to determine the anticipated changes in fit. Static fit testing was completed on FFRs only at NIOSH NPPTL using a static advanced headform (StAH) that quantifies the changes in manikin fit factor. The TSI, Inc. PortaCount® PRO+ 8038 (Shorewood, Minnesota, USA) in “N95 Enabled” mode was used for this evaluation. Assessments of respirators were done using normal and deep breathing without dynamic movements and a speaking passage (11).

### Statistical Analysis

Measurements with standard error bars were generated using Prisma Analytics (Munich, Germany). Means and standard deviations or percent pass of each integrity test method were calculated separately by FFR/MM/CM style. Data for each integrity test method conducted at more than one test site were pooled to create overall means and standard deviations or percent pass. Normality of the data distribution was tested using the Shapiro-Wilk test. Significant differences between untreated and treated FFR/MM/CM were calculated with Student’s t-tests, Mann-Whitney U tests, or Fisher’s exact tests, as appropriate (SAS v9.4). EC_50_s were calculated using GraphPad Prism 8. Full results of integrity tests are presented in Supplemental Table S2A and 2B.

## Supporting information

Supplemental Section

DEMAND STUDY TABLE 1 Expanded

## Data Availability

Data will be freely shared post publication on reasonable request by contacting the corresponding author of the study.

## Acknowledgements

FSKB and YLL acknowledge the donation of masks and respirators from the WHO, as well as the guidance from the WHO CDG, 3M and Halyard. MCC thanks the contribution of the NIOSH National Personal Protective Technology Laboratory: Jonisha P. Pollard, Michael S Bergman, Kevin Strickland, Rebecca Streeter, Christian C. Coby, Nichole Suhon, and Jeremy J Brannen. BHH and MCC acknowledge the biomedical engineering hackathon group through Cassandra Howard and the light boxes built by the Colorado State University Biomedical team, Dr. Jorge Rocca and Han Chi. DE wishes to thank Megan Desaulniers and Dr. Dan Dragon for BSL3 operations at the University of Alberta. JMC acknowledges the assistance of Johanna Blaak and Michelle Wright of the W21C Research and Innovation Centre, University of Calgary for consultation on methodology and for project management, respectively, Jeanette Adams and Rhea Campbell of Alberta Health Services for conducting the human fit testing, and David Silverstone from Alberta Health Services for oversight and management. ET wishes to thank Amélie Matton, Murielle Perrin and Frédéric de Meulemeester (AMB Ecosteryl, Mons, Belgium) for suggestions and technical and administrative support and thanks Chantal Vanmaercke and Carine Boone for their excellent technical support. MCC thanks the administrative support of Mary Moua. TSL acknowledges the support of the Seattle Children’s Hospital Medical Director of Infection Prevention, Dr. Danielle Zerr, and the Chief Medical Officer, Dr. Ruth McDonald.

## Disclaimer

The findings and conclusions in this report are those of the authors and do not necessarily represent the official position of the Centers for Disease Control and Prevention.

## Author contributions

TSL, JC, BHH, FEMS, MCC, YLL, FSKB, DE designed the study methodology. SC provided methodology design and concepts. JC and BH collated and analyzed Methylene Blue clinical chemistry data with NM and OJ developing and performing Methylene Blue test protocols. AP coordinated the Writing and Communications Group. BHH and FEMS coordinated the protocol, standardization and performance of the Virus Study Group (TSL, YL, KBK, EH, ET, YC, TC, CNM, LFLB, DE, JS, CW, TW, JC). SRT, LD, HN, JFW, SDJ contributed to technical operation and generated data needs of the Virus Study Group. TG and ET contributed the MHV methodology and virus stock to the Seattle Children’s Research Institute. FSKB and YLL coordinated the evaluation performance, data validation and standardization of the methodology of the PPE Integrity Performance Group (SJS, AP, RP, MM, YC) with JO, VMS, JLL providing the test methods and data collection for the integrity performance of the masks and respirators. YC coordinated mask and respirator particulate testing by LL and MZ. MML provided the statistical analysis of the study data with assistance by PF for the PortaCount data at the Calgary site. The PortaCount human fit testing for the decontaminated masks at the Calgary site was planned by JMD, KP and SS and coordinated by RJM and JMC with assistance from SR, PF, and KH. AP and LFC provided PortaCount human fit testing data from the (Anesthesia Informatics and Media) AIM lab site. MCC, KH, ML coordinated the DeMaND study administration and logistics. All authors were provided the final manuscript for comments and approval.

## SUPPLEMENTAL FIGURES AND TABLES

- ***TABLE S1:*** Respirator and Mask Decontamination and Testing Matrix
- ***TABLE S2A*:** Integrity Results for Face Masks (MMs and CM)
- ***TABLE S2B*:** Integrity Testing Results for Respirators (FFRs)
- Background: Methylene Blue as a Photosensitizer:
  - ***FIGURE S1***: *Absorption spectrum of a 10 µM methylene blue solution in distilled water*.
  - ***FIGURE S2***: *Photosensitized production of* ^*1*^*O*_*2*_ *by Methylene Blue (MB)*.
  - ***TABLE S3*:** Viral log reductions achieved by MB photodynamic therapy.
- Singlet Oxygen Production Rate - Theoretical Evaluation.
  - ***FIGURE S3***: *Irradiance spectrum of the Husky light source measured at a distance that generates a total illuminance of 100,000 lux*.
  - ***TABLE S4:*** *Calculated fraction of photon absorbed per molecule of MB per second (P) for various light sources*.
  - ***TABLE S5:*** *Total number of photons absorbed per second by the external surface of the mask*.
  - ***TABLE S6:*** *Number of singlet oxygen molecules produced per second on the entire external surface of the mask*.
- ***FIGURE S4:*** Mask Strap Viral Inactivation with MBL or DH
- Tensile Testing of Elastomeric Straps, Ear Loops, and Ties
- Simulated Wear and Air Flow Difference Detection using Sheffield Dummy
  - ***FIGURE S5***: *Effect of MBL, DH, and VHP+O*_*3*_ *treatments on the air flow rate differences measured with the Sheffield Dummy Head for three FFRs (R3, RH, RM), two MMs (FH, FW) and CM during inhalation and exhalation*.
- **Supplemental Materials and Methods**
  - Light boxes
    - ***FIGURE S6:*** *Two constructed light boxes used in the study*
  - Tensile Strength Testing
  - Sheffield AFD Testing

## REFERENCES

1. WHO. Rational use of personal protective equipment for coronavirus disease 2019 (COVID-19). World Health Organization. 2020;6 (April):1–28.

2. United States Food and Drug Administration. Decontamination Systems for Personal Protective Equipment EUAs. 2020;1–4. Available from: https://www.fda.gov/medical-devices/coronavirus-disease-2019-covid-19-emergency-use-authorizations-medical-devices/decontamination-systems-personal-protective-equipment-euas

3. Battelle. Final Report for the Bioquell Hydrogen Peroxide Vapor (HPV) Decontamination for Reuse of N95 Respirators. 2016;46.

4. Costa L, Faustino MAF, Neves MGPMS, Cunha Â, Almeida A. Photodynamic inactivation of mammalian viruses and bacteriophages. Viruses. 2012;4(7):1034–74.

5. Tuite EM, Kelly JM. New trends in photobiology. Photochemical interactions of methylene blue and analogues with DNA and other biological substrates. Journal of Photochemistry and Photobiology, B: Biology. 1993;21(2–3):103–24.

6. Boix-Garriga E, Rodríguez-Amigo B, Planas O, Nonell S. Chapter 2 Properties of Singlet Oxygen. In: Singlet Oxygen: Applications in Biosciences and Nanosciences, Volume 1 [Internet]. The Royal Society of Chemistry; 2016. p. 23–46. Available from: http://dx.doi.org/10.1039/9781782622208-00023

7. Muller-Breitkreutz K, Mohr H, Briviba K, Sies H. Inactivation of viruses by chemically and photochemically generated singlet molecular oxygen. J Photochem Photobiol B [Internet]. 1995/09/01. 1995;30(1):63–70. Available from: https://www.ncbi.nlm.nih.gov/pubmed/8558363

8. Wagner SJ. Virus inactivation in blood components by photoactive phenothiazine dyes. Transfusion Medicine Reviews. 2002;16(1):61–6.

9. Bergman MS, Viscusi DJ, Heimbuch BK, Wander JD, Sambol AR, Shaffer RE. Evaluation of multiple (3-Cycle) decontamination processing for filtering facepiece respirators. Journal of Engineered Fibers and Fabrics. 2010;5(4):33–41.

10. Yan J, Guha S, Hariharan P, Myers M. Modeling the Effectiveness of Respiratory Protective Devices in Reducing Influenza Outbreak. Risk Analysis. 2019;39(3):647–61.

11. NIOSH. Decontaminated Respirator Assessment Plan. NIOSH. 2020. Available from: https://www.cdc.gov/niosh/npptl/respirators/testing/pdfs/NIOSHApproved_Decon_TestPlan10.pdf

12. Kasloff S, Strong J, Funk D, Cutts T. Stability of SARS-CoV-2 on Critical Personal Protective Equipment. medrxiv Jan 1, 2020.

13. Fisher EM. and Shaffer RE. A method to determine the available UV-C dose for the decontamination of filtering facepiece respirators. Journal of applied microbiology 110.1 (2011): 287–295.

14. Fischer RJ, Morris DH, Doremalen N Van Sarchette S, Matson MJ, Bushmaker T, et al. Effectiveness of N95 respirator decontamination and reuse against SARS-CoV-2 Virus. Emerging Infectious Diseases. 2020;26(9):2253–5.

15. Viscusi DJ, King WP, Shaffer RE. Effect of decontamination on the filtration efficiency of two filtering facepiece respirator models. International Society for Respiratory Protection [Internet]. 2007;24(3/4):93. Available from: https://www.isrp.com/the-isrp-journal/journal-public-abstracts/1138-vol-24-no-3-and-no-4-2007-pp-93-107-viscusi-open-access/file

16. Viscusi DJ, Bergman MS, Eimer BC, Shaffer RE. Evaluation of five decontamination methods for filtering facepiece respirators. Annals of Occupational Hygiene. 2009;53(8):815–27.

17. Yan R, Chillrud S, Magadini DL, Yan B. surgical facial masks?: stretching supplies and better protection during the ongoing COVID-19 Pandemic. Journal of the International Society for Respiratory Protection. 2020;37(1):19–35.

18. Price A, Cui Y, Liao L, Xiao W, Chu S, Chu L. Is the fit of N95 facial masks effected by disinfection?? A study of heat and UV disinfection methods using the OSHA protocol fit test. medrxiv Jan 1,2020.

19. Campos RK, Jin J, Rafael GH, Zhao M, Liao L, Simmons G, et al. Decontamination of SARS-CoV-2 and Other RNA Viruses from N95 Level Meltblown Polypropylene Fabric Using Heat under Different Humidities. ACS Nano. 2020;14(10):14017–25.

20. Zhao M, Liao L, Xiao W, Yu X, Wang H, Wang Q, et al. Household materials selection for homemade cloth face coverings and their filtration efficiency enhancement with triboelectric charging. Nano Letters. medrxiv June 2, 2020.

21. CDC. Implementing Filtering Facepiece Respirator (FFR) Reuse, Including Reuse after Decontamination, When There Are Known Shortages of N95 Respirators. CDC. 2020. Available from: https://www.cdc.gov/coronavirus/2019-ncov/hcp/ppe-strategy/decontamination-reuse-respirators.html

22. Rengelshausen J, Burhenne J, Fröhlich M, Tayrouz Y, Singh SK, Riedel KD, et al. Pharmacokinetic interaction of chloroquine and methylene blue combination against malaria. European Journal of Clinical Pharmacology. 2004;60(10):709–15.

23. Krespi YP, Kizhner V. Laser-assisted nasal decolonization of Staphylococcus aureus, including methicillin-resistant Staphylococcus aureus. American Journal of Otolaryngology - Head and Neck Medicine and Surgery [Internet]. 2012;33(5):572–5. Available from: http://dx.doi.org/10.1016/j.amjoto.2012.02.002

24. Degesys NF, Wang RC, Kwan E, Fahimi J, Noble JA, Raven MC. Correlation Between N95 Extended Use and Reuse and Fit Failure in an Emergency Department. JAMA [Internet]. 2020 Jul 7;324(1):94. Available from: https://jamanetwork.com/journals/jama/fullarticle/2767023

25. Boscarino JA, Logan HL, Lacny JJ, Gallagher TM. Envelope protein palmitoylations are crucial for murine coronavirus assembly. Journal of virology [Internet]. 2008 Mar;82(6):2989–99. Available from: http://www.ncbi.nlm.nih.gov/pubmed/18184706

26. Pensaert M, Callebaut P, Vergote J. Isolation of a porcine respiratory, non-enteric coronavirus related to transmissible gastroenteritis. The Veterinary quarterly. 1986;8(3):257–61.

27. Wesley RD, Hill HT, Biwer JD. Evidence for a porcine respiratory coronavirus, antigenically similar to transmissible gastroenteritis virus, in the United States. Vol. 2, Journal of Veterinary Diagnostic Investigation. 1990. p. 312–7.

28. Cox E, Hooyberghs J, Pensaert MB. Sites of replication of a porcine respiratory coronavirus related to transmissible gastroenteritis virus. Research in veterinary science. 1990;48(2):165–9.

29. NIOSH. Determination of Particulate Filter Efficiency Level for N95 Series Filters against Solid Particulates for Non-Powered, Air-Purifying Respirators Standard Testing Procedure (STP). [Internet]. NIOSH. 2019 [cited 2020 Apr 30].

30. NIOSH. 2 CFR Part 84. NIOSH [Internet]. 2020 [cited 2020 Apr 20]; Available from: https://ecfr.io/Title-42/pt42.1.84

31. ASTM. F2100-19e1 Standard Specification for Performance of Materials Used in Medical Face Masks. ASTM International. West Conshohocken, PA; 2020.

32. ASTM. F2101-19 Standard Test Method for Evaluating the Bacterial Filtration Efficiency (BFE) of Medical Face Mask Materials, Using a Biological Aerosol of Staphylococcus aureus. ASTM International. West Conshohocken, PA: ASTM International(2019b); 2019.

33. CEN CWA 17553 Community face coverings - Guide to minimum requirements, methods of testing and use, 2020. https://www.cencenelec.eu/research/CWA/Documents/CWA17553_2020.pdf

34. NIOSH. Determination of Exhalation Resistance Test, Air-Purifying Respirators Standard Testing Procedure (STP) [Internet]. NIOSH 2019. Available from: https://www.cdc.gov/niosh/npptl/stps/pdfs/TEB-APR-STP-0003-508.pdf

