## Supplemental Section for "Addressing Personal Protective Equipment (PPE) Decontamination: Methylene Blue and Light Inactivates SARS-CoV-2 on N95 Respirators and Masks with Maintenance of Integrity and Fit"

### SUPPLEMENTAL MATERIAL

### Background: Methylene Blue as a Photosensitizer

Methylene blue (MB) is a positively charged phenothiazinium dye first synthesized in 1876 for use as a textile dye (1). The blue color of MB results from a strong absorption band in the red visible light spectrum with a peak at 664 nanometers as illustrated in [FIGURE S1]. ($\varepsilon_{664}$ = 85,000 $M^{-1}$ $cm^{-1}$) (2). Absorption of light by MB generates singlet oxygen ($^{1}O_2$) and other reactive species, which are microbicidal, a process known as photodynamic therapy (1,)2). The initial step corresponds to the excitation of MB into its singlet excited state upon absorption of a photon, which is followed by an efficient intersystem crossing to the molecule's triplet excited state (triplet quantum yield, $\Phi_T = 0.52$) (2). The latter generates singlet oxygen almost quantitively via energy transfer (singlet oxygen quantum yield $\Phi_{\Delta} \sim 0.50$) (3). Alternatively, a small portion of the triplet excited state is deactivated via electron transfer to a reducing agent, which leads ultimately to the formation of a superoxide anion [FIGURE S2] (2).

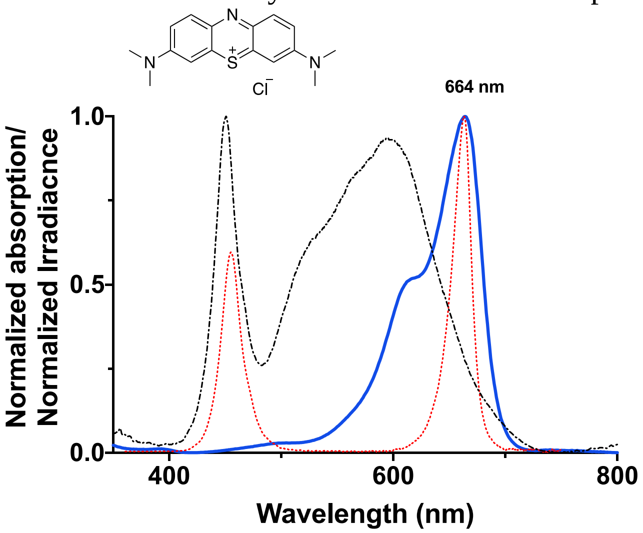

*__FIGURE S1:__ Normalized absorption spectrum of a 10 μM solution of methylene blue (MB) (blue trace) in DI water recorded using a Cary 50 spectrophotometer (Agilent) using a quartz cuvette (1cm pathlength) with an overlay of the normalized irradiance spectrum of each light source used in this study. The Husky lamp irradiance spectrum ($W/m^2$, black dashed trace) was recorded using a Stellar-RAD spectroradiometer (StellarNet Inc.); whereas the red lamp irradiance spectrum ($W/m^2$, red dotted trace) was measured via a EKO Wiser spectroradiometer equipped with a MS-711 sensor. The chemical structure of MB is shown at the top of the figure.*

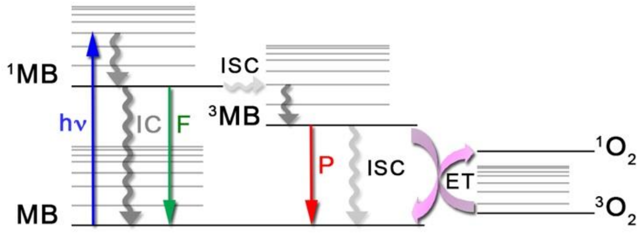

***FIGURE S2: Photosensitized production of $^{1}O_2$ by Methylene Blue (MB).*** *Upon absorption of a photon (hν), methylene blue (MB) is promoted to one the of the vibrational levels of the molecule's singlet excited state. The latter deactivates rapidly and returns to the lowest vibrational level of the first excited state ($^{1}$MB). From there, the excited molecule will lose its energy by a combination of both radiative and non-radiative processes. IC*

*corresponds to internal conversion, a non-radiative process, while F refers to fluorescence, a radiative process and ISC stands for intersystem crossing, another non-radiative process. Once $^{1}MB$ intersystem cross to its triplet excited state ($^{3}MB$), it can go back to the ground state non-radiatively via intersystem crossing or radiatively by emission of light referred to as phosphorescence (P). Alternatively, $^{3}MB$ can transfer its energy to surrounding molecular oxygen ($^{3}O_2$) resulting in the formation of singlet oxygen ($^{1}O_2$).*

Singlet oxygen is the lowest excited state of molecular oxygen and can be viewed as an "energized" form of oxygen (4). Because it is an excited state species, singlet oxygen has a finite lifetime, which is environment-dependent. In water, singlet oxygen has a very short lifetime of ~3 μs (4) while in ambient air it can last up to 90 ms, thus creating different spheres of reactivity from hundreds of nanometers in aqueous environment to a few millimeters in air (2,3,5). These different lifetimes translate into different diffusion lengths providing singlet oxygen with different spheres of reactivity ranging from hundreds of nanometers in aqueous environment to a few millimeters in air (4,5). However, if left unreacted, singlet oxygen simply reverts back to molecular oxygen (4). On the other hand, the reaction of singlet oxygen with biomolecules can lead to formation of secondary reactive species extending the microcidal effects of photodynamic therapy (6,7).

MB, without light activation, is approved by the U.S. Food and Drug Administration for treatment of acquired methemoglobinemia in children and adults by intravenous injection, at a dose of 1 mg/kg bodyweight followed by a second similar dose as needed (US FDA). Topical application of MB in the mouth or nose followed by light activation is approved for use as a photodynamic microbicidal therapy for periodontitis in Canada, the United Kingdom, and the European Union, and for pre-operative intranasal disinfection in Canada (8,9). MB and light is also routinely used commercially in Europe, South America, and Asia Pacific regions for viral disinfection of pooled human donor plasma (10). MB is listed as one of the WHO's Model List of Essential Medicines 2019 (11). MB is also widely used in a variety of off label indications including:

- Identification of lymph nodes for sentinel node biopsy after subcutaneous injection (12)
- As a marker dye for Barrett's esophagus and colonic polyps during endoscopic evaluation (13,14)
- As an injection to identify ureteral leakage from trauma and the existence of pelvic fistulas (15,16)
- Treatment of ifosfamide neurotoxicity and cyanide poisoning (17)
- To evaluate aspiration of oral contents into the lung (18)
- In clinical trials as a vasopressor for septic shock, as cognitive enhancer for Alzheimer's disease, as an oral treatment for malaria, and as a topical treatment for hidradenitis suppurativa (17)

One of the first studies demonstrating an antiviral effect of MB and light against a herpes virus was published in 1932 by Perdrau (19). Subsequent mechanisms of action research has shown that the photodynamic generation of singlet oxygen by light activated MB attacks lipids, proteins, and nucleic acids, and has been demonstrated in multiple studies to inactivate a range of DNA and RNA viruses, including Ebola virus, Middle East Respiratory Syndrome Coronavirus, and more recently, SARS-CoV-2 (20–22) [TABLE S3].

***TABLE S3. Viral log reductions achieved by MB photodynamic therapy.***

| *Reference* | Pathogen | Log Reduction |
|---|---|---|
| (10) | | |
| | *Enveloped viruses* | |
| | Human immunodeficiency virus-1 (HIV-1) | ≥5.45 |
| | West Nile Virus (WNV) | ≥5.78 |
| | Bovine viral diarrhea virus (BVDV) | ≥5.44 |
| | Pseudorabies virus (PRV) | ≥5.48 |
| | Duck hepatitis B virus (HBV) | ≥6 |
| | Influenza H3N2 | ≥4.40 |
| | Cytomegalovirus (CMV) | ≥4.08 |
| | Infectious bronchitis virus (IBV) | ≥4.90 |
| | Hog cholera | ≥5.92 |
| | Herpes simplex virus | ≥5.50 |
| | Bovine herpes | ≥8.11 |
| | Semliki Forest virus | ≥7.00 |
| | Sindbis virus | ≥9.73 |
| | Influenza virus | ≥5.1 |
| | Vesicular stomatitis virus (VSV) | ≥4.89 |
| | *Non-enveloped viruses* | |
| | Human adenovirus-5 (Had-5) | ≥5.33 |
| | Calicivirus | ≥3.9 |
| | Simian virus 40 (SV40) | ≥4 |
| | Parvovirus B19 | ≥5 |
| | Porcine parvovirus | ≥0 |
| | Poliovirus | ≥1 |
| | Hepatitis A | ≥0 |
| (23) | | |
| | Ebola | ≥4.7 (light dose 30 J/cm$^2$) |
| | Middle East respiratory syndrome coronavirus (MERS-CoV) | ≥3.3 (light dose 30 J/cm$^2$) |
| (24) | | |
| | Severe acute respiratory syndrome coronavirus (SARS-CoV) | ≥3.1 (light dose 30 J/cm$^2$) |
| | Crimean–Congo hemorrhagic fever virus (CCHFV) | ≥3.2 (light dose 30 J/cm$^2$) |
| | Nipah virus (NiV) | ≥2.7 (light dose 30 J/cm$^2$) |

**Singlet Oxygen Production Rate - Theoretical Evaluation.**

The fraction of photon (P) absorbed by one molecule per unit of time is given by the following integrated product:

$$P = \int_{\lambda_1}^{\lambda_2} f(\lambda)\, \sigma(\lambda)\; d\lambda \qquad \text{(eq. 1)}$$

where f(λ) is the photon flux (#photon/$cm^2$/s) and σ(λ) corresponds to the absorption cross-section of the molecule ($cm^2$).

The photon flux, f(λ), is obtained from the irradiance spectrum of the light source, as the irradiance I(λ) in W/$cm^2$ (= J/s/$cm^2$) corresponds to:

$$I(\lambda) = f(\lambda) \times E(\lambda) \quad \text{(eq 2)}$$

where E(λ) is the energy in Joules (J) of a photon of wavelength λ.

The irradiance spectrum of the Husky lamp was recorded using a Stellar-RAD spectroradiometer (StellarNet Inc.) and is shown in [FIGURE S3]. In this figure, the spectroradiometer was located at a distance where the total illuminance was 100,000 lux.

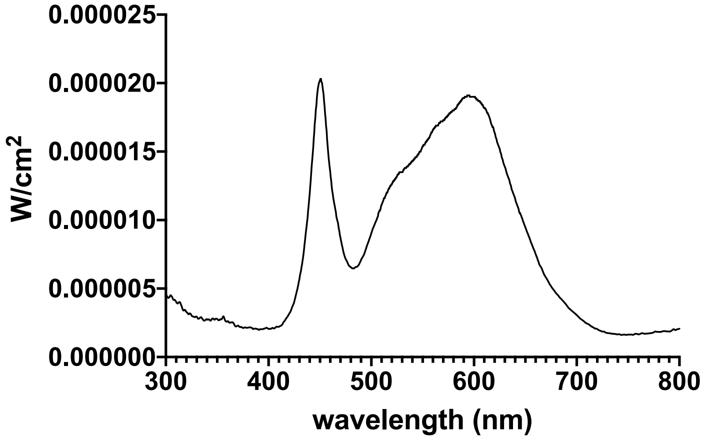

***FIGURE S3: Irradiance spectrum of the Husky light source measured at a distance that generates a total illuminance of 100,000 lux.***

Similarly, the irradiance spectrum of the Husky light source was also recorded when the spectroradiometer was located at a distance where the total illuminance was 50,000 lux. Finally, the irradiance spectrum of fluorescent light bulbs was measured in a room under normal lighting condition at a total illuminance of 1300 lux.

The absorption cross section of one molecule of MB, σ(λ), is obtained from the knowledge of the molecule's molar absorption coefficient, ε(λ), as per equation 3:

$$\sigma(\lambda) = \frac{2.3\times10^3}{N_A}\, \varepsilon(\lambda) = 3.824 \times 10^{-21}\, \varepsilon(\lambda) \quad \text{(eq. 3)}$$

Therefore, under our experimental conditions, the fraction of photon (P) absorbed by one molecule of MB in 1 second is calculated to be 0.86 when the light source is a Husky lamp with a total illuminance of 100,000 lux. The fraction of photon (P) absorbed by one molecule of MB in 1 second becomes 0.56 when the total illuminance of the Husky lamp is reduced to 50,000 lux; whereas it is calculated to be 0.2 under ambient light (fluorescent light source) conditions with a total illuminance of 1300 lux. These data are summarized in [TABLE S4].

***TABLE S4*****:** Calculated fraction of photon absorbed per molecule of MB per second (P) for various light sources.

| | Husky LED (100,000 lux) | Husky LED (50,000 lux) | Fluorescent light (1300 lux) |
|---|---|---|---|
| P | 0.86 | 0.56 | 0.2 |

The number of molecules of MB present on the total external surface of the mask can be obtained from knowledge of the initial concentration of MB and the total volume sprayed. If a total volume of 8 ml of MB solution is applied onto the surface of the mask in 6 sprays, 4 on the outside and 2 on the inside, then 5.33 ml are used to cover the outside surface of the mask. Therefore, $3.21\times10^{16}$ molecules of MB will be present on the external surface of the mask for an initial concentration in MB of 10μM, while this number reduces to $3.21\times10^{15}$ molecules for an initial concentration of MB of 1μM.

If we assume that all photons emitted by the light source are absorbed by the surface of the mask, then by taking into consideration the number of molecules of MB present on the external surface of the mask and the fraction of photon absorbed per molecule per second, the total number of photons absorbed per second can easily be obtained. The latter are summarized in [TABLE S5] for the different light sources.

***TABLE S5:*** Total number of photons absorbed per second by the external surface of the mask.

| | Husky LED (100,000 lux) | Husky LED (50,000 lux) | Fluorescent light (1300 lux) |
|---|---|---|---|
| [MB] = 10μM | $2.8\times10^{16}$ | $1.8\times10^{16}$ | $6.4\times10^{15}$ |
| [MB] = 1μM | $2.8\times10^{15}$ | $1.8\times10^{15}$ | $6.4\times10^{14}$ |

Since the singlet oxygen quantum yield ($\Phi_\Delta$) represents the number of singlet oxygen molecule generated per number of photons absorbed by the photosensitizer, and MB has a $\Phi_\Delta$ of 0.5, then it is estimated that at least $3.2\times10^{14}$ of singlet oxygen molecules are produced per second for the entire external surface of the mask under ambient light irradiation. The number of singlet oxygen molecules produced per second under different irradiation conditions are summarized in [TABLE S6].

***TABLE S6:*** Number of singlet oxygen molecules produced per second on the entire external surface of the mask.

| | Husky LED (100,000 lux) | Husky LED (50,000 lux) | Fluorescent light (1300 lux) |
|---|---|---|---|
| [MB] = 10μM | $1.4\times10^{16}$ | $9\times10^{15}$ | $3.2\times10^{15}$ |
| [MB] = 1μM | $1.4\times10^{15}$ | $9\times10^{14}$ | $3.2\times10^{14}$ |

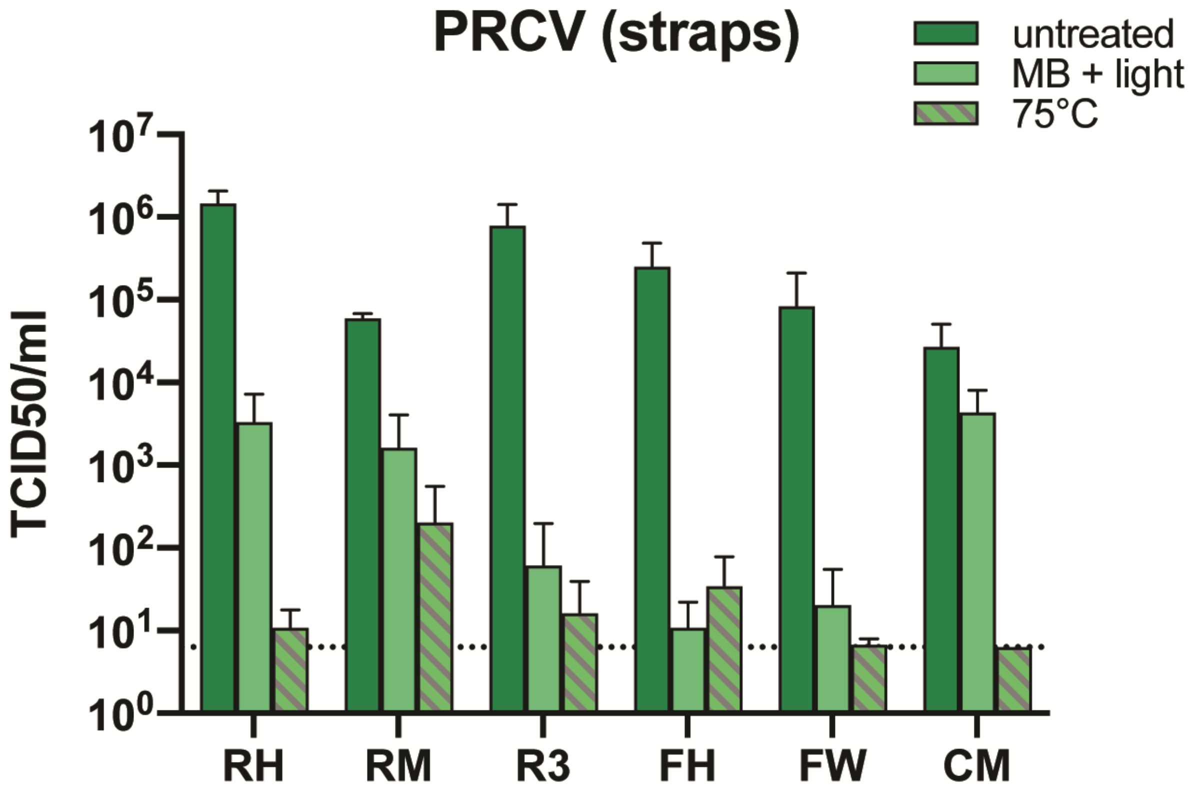

***FIGURE S4. PRCV Inactivation on Inoculated Mask Straps by MBL or DH.***

***(A)*** *100 µl of PRCV was added onto both straps of FFRs, MMs and a CM. The straps were allowed to dry for 30 minutes before exposure to dry heat (60 min at 75°C) or 10 µM MB + light (30 min). Six inoculated, decontaminated straps (n=6) were analyzed in parallel to inoculated, untreated, positive control straps (n=6). Subsequently viral titers were determined by* $TCID_{50}$ *assay. Data is represented as mean +/- SD. CM= homemade community mask. FH= Type IIR Halyard face mask. FW= Type II generic face mask. RH= Halyard duckbill respirator. FW= Type II generic face mask. R3= 3M panel respirator (1870+). Dotted line represents the lower limit of detection.*

**Tensile Testing of Elastomeric Straps, Ear Loops, and Ties**

The changes in the elastic recovery of the FFR straps and MM ear loops were examined to determine changes in strap integrity due to the decontamination process, using a method defined in the National Personal Protection Technology Laboratory (NPPTL) Decontaminated Respirator Assessment Plan (25). The straps of the VHP+$O_3$ treated RMs were frayed in several locations and significant deterioration was observed. This was also reflected in the human fit testing. Straps of RMs also showed increases in force in both the top and bottom straps after VHP+$O_3$. No visual degradation of the straps of any other model/decontamination method was observed. Inconsistent changes were shown between the top and bottom straps with MBL-treated RM. The RM (DH) and RH (MBL, DH, and VHP+$O_3$) showed decreases in recorded force for both the top and bottom straps. The R3 (MBL, DH, and VHP) showed increases in recorded force for both the top and bottom straps.

Also, all three decontamination methods significantly impacted the elastic recovery of the ear loops of the FHs while VHP treatment significantly reduced both MMs ($p<0.01$). The tensile strength and elongation of the CM ties were tested using a different method (EN ISO 13934-1:2013) because the ties were made of non-elastic folded and sewn fabrics. Statistical analysis revealed significant changes on the tensile strength of CM ties. After MBL treatment it was observed that the elongation was significantly increased. However, since these masks were made by the community, a variability in the fabric properties was expected and noted.

**Simulated Wear and Air Flow Difference Detection using Sheffield Dummy**

Simulated wear air flow differences (SW-AFD) using a Sheffield dummy head connected to a breathing circuit were used to mimic rates of breathing ranging from shallow (28.3 L/min) to deep breathing (85 L/min and 160 L/min). The maximum air flows detected were similar to pressure drop differences, but tested at four air flow rates. Note that inhalation and exhalation AFDs at 85 L/min, Figure S5b and S5c, were similar in values. At higher air flows AFDs were also higher. As seen in the FIGURE S5, MBL treatment did not alter AFD with inhales and exhales. DH treatments significantly impacted AFD of RM and R3 FFRs compared to untreated for inhalation at 28.3 L/min and 85 L/min, and exhalation at 85 L/min.

Significant differences were detected between DH-treated and control FFR samples for RM and panel R3 shapes, for all airflows except the highest exhalation flow of 160 L/min. At shallow inhalation (28.3 L/min), DH-RM AFD were significantly lower compared to untreated RM (-0.21 L/min compared to -0.26 L/min) and DH-R3 AFD were higher compared to untreated R3 (-0.25 L/min versus -0.22 L/min). At medium inhalation (85 L/min), DH-RM AFD was significantly lower compared to untreated RM (- 0.61 L/min versus -0.79L/min) and DH-R3 AFD was significantly higher compared to untreated R3 (-0.75 L/min versus -0.66L/min). At medium exhalation (85 L/min), DH-RM AFD was significantly lower compared untreated RM (0.59 L/min versus 0.74 L/min) and DH-R3 AFD was significantly higher compared to untreated R3 (0.73L/min versus 0.63 L/min).

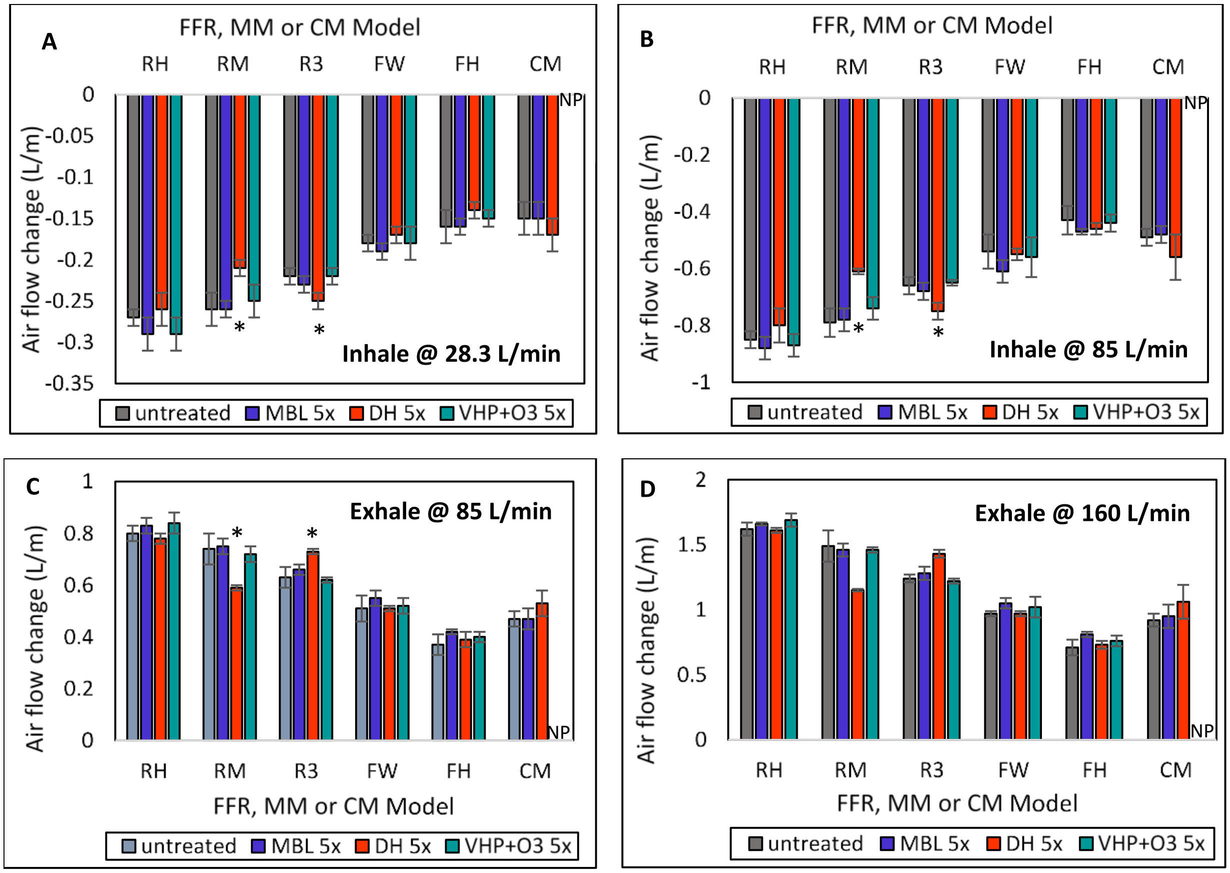

***Figure S5. Effect of MBL, DH, and VHP+O$_3$ treatments on the air flow rate differences measured with the Sheffield Dummy Head for three FFRs (R3, RH, RM), two MMs (FH, FW) and CM during inhalation and exhalation. (A)****: Inhale 28.3 L/m Test* ***(B)*** *Inhale 85 L/m Test.* ***(C)*** *Exhale 85 L/m Test.* ***(D)*** *Exhale 160 L/m Test. Air flow rate changes were used to mimic rates of breathing ranging from shallow to deep breathing. RH= Halyard duckbill respirator (Fluidshield-46727). RM= 3M half-sphere respirator (1860). R3= 3M panel respirator (1870+). FW= EN 14683 Type II generic face mask. FH= ASTM F2100 Level 2 Halyard face mask. CM=Community mask. * Results from decontaminated respirators and masks are significantly different from untreated respirators and masks (Student's t test or Mann-Whitney U test $p < 0.05$).*

### Supplemental Materials and Methods

#### **Light Boxes**

The illumination system created at Colorado State University and used at Seattle Children's Hospital, University of Calgary, George Washington University, and Nelson Laboratories consists of a transparent acrylic shelf placed horizontally to support the objects to be disinfected, surrounded by two Husky LED light panels (model# K40187) on top and bottom [FIGURE S6].

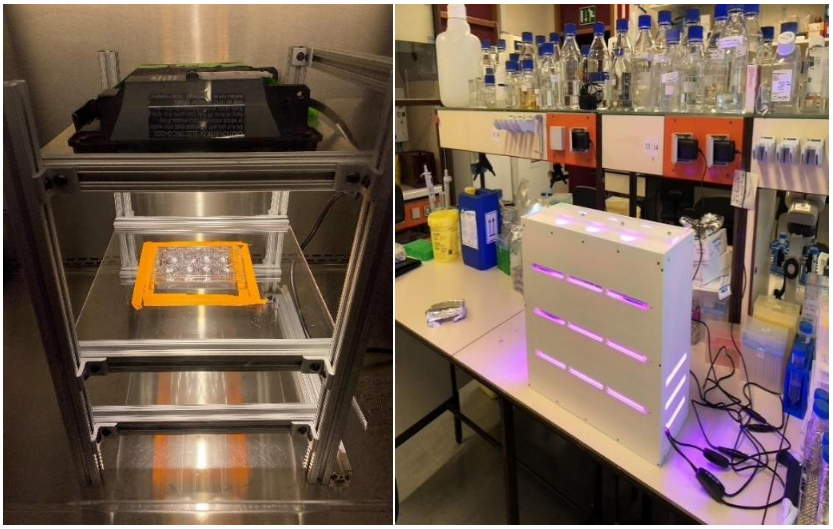

***FIGURE S6. Two constructed light boxes used in the study. Left:*** *Husky Light Box. This light box designed and constructed by the Hackathon Team at Colorado State University is configured with one Husky LED light. The clear acrylic shelf is positioned at a distance where the Husky light applies 50,000 lux of illuminance to the area delineated by the orange tape as measured by a light meter. This light box can also be fitted with a second Husky LED at the bottom to apply light to both sides of masks or respirators placed on the shelf.* ***Right:*** *University of Liege designed light box. This light box contains 3 x 6 Roleadro 60W Culture Indoor IP65 LED Horticultural T5 Grow Lamps with dimmable light control providing red light (610-720 nm) and blue light (400-520 nm) spectrum of light.*

The distance between the light panels and the objects could be adjusted by displacing the shelve on the aluminum rails that comprise the supporting frame of the light box. Each of the two 4000 K color temperature LED light panels consumed about 56 W of electric power and provided a white light power of 13.7 W measured at the illumination surface of the LEDs. The spectrally integrated band between 560-710 nm of light coming out of the LED panel surface band had a divergence leading to a power density distribution ranging from about 170 $W/m^2$ at the center to 90 $W/m^2$ at the edge of a 10" (25.4 cm) diameter circular at 6" (15 cm) from the LED panel surface. This corresponded to an in-band illuminance of 64,300 and 34,000 lux (lux= lumen/m2), respectively. Similar Husky LEDs were used at the University of Alberta but had a 3500K color temperature. A digital light meter (Latnex LM-50KL) and shims were used to identify locations where the illumination within the bio containment hood equaled 50,000 lux. Light intensity was determined using a light meter (Latnex, model LM-50KL or Cooke, Model CK-CL400). The University of Liege and Centexbel used a horticultural light box for their studies containing 3 x 6 red/blue 60 W horticultural LED lamps (Roleadro Culture Indoor IP65 LED Horticultural T5 Grow). Light intensity was determined using a light meter (DeltaOHM, Model HD2102.2).

**Tensile Strength Testing**

The changes in the elastic recovery of the respirator straps and mask ear loops was performed to determine changes in strap integrity due to the decontamination process using a method defined in NIOSH Decontaminated Respirator Assessment Plan (26). The tensile strength and elongation of the CM ties were tested using a different method (EN ISO 13934-1:2013) because the ties were made of non-elastic folded and sewn fabrics.

**Sheffield AFD Testing**

The simulated wear treatment set-up of clause 7.16 of the EN 149 standard was used to determine differences in air flow between untreated and decontaminated respirators and masks. Three simulated wear conditions mimicked shallow to deep breathing and at variations of conventional flow rates: inhalation at 28.3 L/min (EN 149 and EN 14683), inhalation at 85 L/min (NIOSH N95) and exhalation at 85 L/min (NIOSH N95) and exhalation at 160 L/min (EN 149). Since this modified method is not a requirement for mask or respirator certification, we used Sheffield AFD to detect differences caused by the decontamination process compared to untreated masks or respirators.

### References

1. Wainwright M, Crossley KB. Methylene Blue - a Therapeutic Dye for All Seasons? Journal of Chemotherapy [Internet]. 2002;14(5):431–43. Available from: https://doi.org/10.1179/joc.2002.14.5.431
2. Tardivo JP, Del Giglio A, de Oliveira CS, Gabrielli DS, Junqueira HC, Tada DB, et al. Methylene blue in photodynamic therapy: From basic mechanisms to clinical applications. Photodiagnosis and Photodynamic Therapy [Internet]. 2005;2(3):175–91. Available from: http://www.sciencedirect.com/science/article/pii/S1572100005000979
3. Fernandez JM, Bilgin MD, Grossweiner LI. Singlet oxygen generation by photodynamic agents. Journal of Photochemistry and Photobiology B: Biology [Internet]. 1997;37(1):131–40. Available from: http://www.sciencedirect.com/science/article/pii/S1011134496073496
4. Boix-Garriga E, Rodríguez-Amigo B, Planas O, Nonell S. Chapter 2 Properties of Singlet Oxygen. In: Singlet Oxygen: Applications in Biosciences and Nanosciences, Volume 1 [Internet]. The Royal Society of Chemistry; 2016. p. 23–46. Available from: http://dx.doi.org/10.1039/9781782622208-00023
5. Midden WR, Wang SY. Singlet oxygen generation for solution kinetics: clean and simple. Journal of the American Chemical Society [Internet]. 1983;105(13):4129–35. Available from: https://doi.org/10.1021/ja00351a001
6. Ouédraogo GD, Redmond RW. Secondary Reactive Oxygen Species Extend the Range of Photosensitization Effects in Cells: DNA Damage Produced Via Initial Membrane Photosensitization¶†. Photochemistry and Photobiology [Internet]. 2003;77(2):192–203. Available from: https://doi.org/10.1562/0031-8655(2003)0770192SROSET2.0.CO2
7. Redmond RW, Kochevar IE. Spatially Resolved Cellular Responses to Singlet Oxygen. Photochemistry and Photobiology [Internet]. 2006;82(5):1178. Available from: https://doi.org/10.1562/2006-04-14-IR-874
8. Branson RD, Blakeman TC, Robinson BRH, Johannigman JA. Use of a single ventilator to support 4 patients: Laboratory evaluation of a limited concept. Respiratory Care. 2012;57(3):399–403.
9. Joseph B, Janam P, Narayanan S, Anil S. Is antimicrobial photodynamic therapy effective as an adjunct to scaling and root planing in patients with chronic periodontitis? A systematic review. Biomolecules. 2017;7(4):79.
10. Seghatchian J, Walker WH, Reichenber S: Updates on pathogen inactivation of plasma using Theraflex methylene blue system. Transfus Apher Sci 2008; 38:271–80.
11. Organización Mundial de la Salud (OMS). World health organization model list of essential medicines. Mental and Holistic Health: Some International Perspectives. 2019;21:119–34.
12. Li J, Chen X, Qi M, Li Y. Sentinel lymph node biopsy mapped with methylene blue dye alone in patients with breast cancer: A systematic review and metaanalysis. PLoS ONE. 2018;13(9):1–18.
13. Canto MI. Vital staining and Barrett's esophagus. Gastrointestinal Endoscopy. 1999;49(3 II).
14. Buchner AM. The Role of Chromoendoscopy in Evaluating Colorectal Dysplasia. Gastroenterology & Hepatology. 2017;13(6):336–47.
15. More R. What is the role of f indigo carmine or methylene blue in the intraoperative

workup of ureteral injury ? MedScape. 2018;9–12.
16. Hanash KA, Al Zahrani H, Mokhtar AA, Aslam M. Retrograde Vaginal Methylene Blue Injection for Localization of Complex Urinary Fistulas. Journal of Endourology. 2003;17(10):941–3.
17. Koch R. Some Drugs and Herbal Products. IARC monographs on the evaluation of carcinogenic risks to humans. 2016;108:7–419.
18. Garuti G, Reverberi C, Briganti A, Massobrio M, Lombardi F, Lusuardi M. Swallowing disorders in tracheostomised patients: A multidisciplinary/multiprofessional approach in decannulation protocols. Multidisciplinary Respiratory Medicine. 2014;9(1):1–10.
19. Virus THE, Immunised H, Virus L. Rabbits immunised with living virus. National Institute for Medical Research. 1935;(392):447–55.
20. Eggers M, Koburger-Janssen T, Eickmann M, Zorn J. In Vitro Bactericidal and Virucidal Efficacy of Povidone-Iodine Gargle/Mouthwash Against Respiratory and Oral Tract Pathogens. Infectious Diseases and Therapy [Internet]. 2018;7(2):249–59. Available from: https://doi.org/10.1007/s40121-018-0200-7
21. Muller-Breitkreutz K, Mohr H, Briviba K, Sies H. Inactivation of viruses by chemically and photochemically generated singlet molecular oxygen. J Photochem Photobiol B [Internet]. 1995/09/01. 1995;30(1):63–70. Available from: https://www.ncbi.nlm.nih.gov/pubmed/8558363
22. Yan J, Guha S, Hariharan P, Myers M. Modeling the Effectiveness of Respiratory Protective Devices in Reducing Influenza Outbreak. Risk Analysis. 2019;39(3):647–61.
23. Eickmann M, Gravemann U, Handke W, Tolksdorf F, Reichenberg S, Müller TH, et al. Inactivation of Ebola virus and Middle East respiratory syndrome coronavirus in platelet concentrates and plasma by ultraviolet C light and methylene blue plus visible light, respectively. Transfusion. 2018;58(9):2202–7.
24. Eickmann M, Gravemann U, Handke W, Tolksdorf F, Reichenberg S, Müller TH, et al. Inactivation of three emerging viruses – severe acute respiratory syndrome coronavirus, Crimean–Congo haemorrhagic fever virus and Nipah virus – in platelet concentrates by ultraviolet C light and in plasma by methylene blue plus visible light. Vox Sanguinis. 2020;115(3):146–51.
25. NIOSH. Decontaminated Respirator Assessment Plan. NIOSH (2020a). 2020. Accessed November 26, 2020 at https://www.cdc.gov/niosh/npptl/respirators/testing/pdfs/NIOSHApproved_Decon_TestPlan10.pdf
26. NIOSH. Determination of Particulate Filter Efficiency Level for N95 Series Filters against Solid Particulates for Non-Powered, Air-Purifying Respirators Standard Testing Procedure (STP). [Internet]. NIOSH (2019a). 2019 [cited 2020 Apr 30]. Available from: https://www.cdc.gov/niosh/npptl/stps/pdfs/TEB-APR-STP-0059-508.pdf
