## Supplemental Table S1 for "Addressing Personal Protective Equipment (PPE) Decontamination: Methylene Blue and Light Inactivates SARS-CoV-2 on N95 Respirators and Masks with Maintenance of Integrity and Fit"

### Table S1: Respirator and Mask Decontamination and Testing Matrix

| **Viricidal Efficacy** | | | **Respirator or Mask Type:** | **Halyard Duckbill Respirator (RH)** | **3M 1860 Half-sphere Respirator (RM)** | **3M 1870+ Panel Respirator (R3)** | | | **WHO Type II Face Mask (FW)** | **Halyard Type IIR Face Mask (FH)** | **Community Face Mask (CM)** |
|---|---|---|---|---|---|---|---|---|---|---|---|
| **Decontamination Site** | **Decontamination Method** | **Respirator Virucidal Test** | **Respirator Test site** | | | | **Mask Virucidal Test** | **Mask Test Site** | | | |
| N/A | Untreated after MHV application | Virucidal efficacy on partial mask | University of Washington / Seattle Children's Hospital | 3 | 3 | 3 | Virucidal efficacy on partial mask | University of Washington / Seattle Children's Hospital | 3 | 3 | 3 |
| University of Washington / Seattle Children's Hospital | MBL and DH after MHV application | Virucidal efficacy on partial mask | University of Washington / Seattle Children's Hospital | 3 | 3 | 3 | Virucidal efficacy on partial mask | University of Washington / Seattle Children's Hospital | 3 | 3 | 3 |
| N/A | Untreated after SARS-CoV-2 application | Virucidal efficacy on partial mask | George Washington University | 3 | 3 | 3 | Virucidal efficacy on partial mask | George Washington University | 3 | 3 | 3 |
| George Washington University | MBL and DH after SARS-CoV-2 application | Virucidal efficacy on whole mask | George Washington University | 3 | 3 | 3 | Virucidal efficacy on whole mask | George Washington University | 3 | 3 | 3 |
| N/A | Untreated after SARS-CoV-2 application | Virucidal efficacy on partial mask | University of Alberta | 3 | 3 | 3 | Virucidal efficacy on partial mask | University of Alberta | 3 | 3 | 3 |
| University of Alberta | MBL and DH after SARS-CoV-2 application | Virucidal efficacy on partial mask | University of Alberta | 3 | 3 | 3 | Virucidal efficacy on partial mask | University of Alberta | 3 | 3 | 3 |
| N/A | Untreated after PRCV application | Virucidal efficacy on partial mask | University of Liège | 3 | 3 | 33 | Virucidal efficacy on partial mask | University of Liège | 31 | 3 | 3 |
| University of Liège | MBL after PRCV application | Virucidal efficacy on partial mask | University of Liège | 3 | 3 | 3 | Virucidal efficacy on partial mask | University of Liège | 3 | 3 | 3 |
| University of Liège | DH after PRCV application | Virucidal efficacy on partial mask | University of Liège | 3 | 3 | 3 | Virucidal efficacy on partial mask | University of Liège | 3 | 3 | 3 |
| | | | **Virucidal Efficacy Subtotal:** | **27** | **27** | **57** | | | **55** | **27** | **27** |
| **Integrity Testing** | | | | | | | | | | | |
| **Decontamination Site** | **Decontamination Method** | **Respirator Integrity Test** | **Respirator Test Site** | | | | **Mask Integrity Test** | **Mask Test Site** | | | |
| N/A | untreated | N/A | N/A | N/A | N/A | N/A | Bacterial Filtration Efficiency and Pressure Drop; Fluid Penetration; Tensile Strength | Centexbel | 20 | 20 | 20 |

| | | | | | | | | | | | |
|---|---|---|---|---|---|---|---|---|---|---|---|
| University of Liège | MBL 5x | N/A | N/A | N/A | N/A | N/A | Bacterial Filtration Efficiency and Pressure Drop; Fluid Penetration; Tensile Strength | Centexbel | 20 | 20 | 20 |
| University of Liège | DH 5x | N/A | N/A | N/A | N/A | N/A | Bacterial Filtration Efficiency and Pressure Drop; Fluid Penetration; Tensile Strength | Centexbel | 20 | 20 | 20 |
| N/A | untreated | Human Fit (PortaCount) | University of Calgary | 5 | 5 | 5 | N/A | N/A | N/A | N/A | N/A |
| University of Calgary/ Alberta Health Services | MBL 5x | Human Fit (PortaCount) | University of Calgary | 5 | 5 | 5 | N/A | N/A | N/A | N/A | N/A |
| University of Calgary/ Alberta Health Services | DH 5x | Human Fit (PortaCount) | University of Calgary | 5 | 5 | 5 | N/A | N/A | N/A | N/A | N/A |
| University of Calgary/ Alberta Health Services | VHP+$O_3$ 5x | Human Fit (PortaCount) | University of Calgary | 5 | 5 | 5 | N/A | N/A | N/A | N/A | N/A |
| N/A | untreated | NaCl Filtration Efficiency and Breathing Resistance; Advanced Headform (Manikin); Tensile Strength (respirators and masks) | NIOSH NPPTL | 10 | 10 | 10 | NaCl Filtration and Breathing Resistance; Bacterial Filtration Efficiency and Pressure Drop; Fluid Penetration | Nelson Laboratories | 20 | 20 | 20 |
| Nelson Laboratories | MBL 5x | NaCl Filtration Efficiency and Breathing Resistance; Advanced Headform (Manikin); Tensile Strength (respirators and masks) | NIOSH NPPTL | 13 | 13 | 13 | NaCl Filtration and Breathing Resistance; Bacterial Filtration Efficiency and Pressure Drop; Fluid Penetration | Nelson Laboratories | 20 | 20 | 20 |
| 4C Air Inc. | DH 5x | NaCl Filtration Efficiency and Breathing Resistance; Advanced Headform (Manikin); Tensile Strength (respirators and masks) | NIOSH NPPTL | 13 | 13 | 13 | NaCl Filtration and Breathing Resistance; Bacterial Filtration Efficiency and Pressure Drop; Fluid Penetration | Nelson Laboratories | 20 | 20 | 20 |
| Stryker Ltd. | VHP+$O_3$ 5x | NaCl Filtration Efficiency and Breathing Resistance; Advanced Headform (Manikin); Tensile Strength (respirators and masks) | NIOSH NPPTL | 13 | 13 | 13 | NaCl Filtration and Breathing Resistance; Bacterial Filtration Efficiency and Pressure Drop; Fluid Penetration | Nelson Laboratories | 20 | 20 | N/A |
| N/A | untreated | Sheffield Dummy Fit | BSI LLC | 3 | 3 | 3 | Sheffield Dummy Fit | BSI LLC | 3 | 3 | 3 |

| | | | | | | | | | | | |
|---|---|---|---|---|---|---|---|---|---|---|---|
| Nelson Laboratories | MBL 5x | Sheffield Dummy Fit | BSI LLC | 3 | 3 | 3 | Sheffield Dummy Fit | BSI LLC | 3 | 3 | 3 |
| 4C Air Inc. | DH 5x | Sheffield Dummy Fit | BSI LLC | 3 | 3 | 3 | Sheffield Dummy Fit | BSI LLC | 3 | 3 | 3 |
| Stryker Ltd. | VHP+$O_3$ 5x | Sheffield Dummy Fit | BSI LLC | 3 | 3 | 3 | Sheffield Dummy Fit | BSI LLC | 3 | 3 | N/A |
| N/A | untreated | Human Fit (PortaCount) | Stanford University | 5 | 5 | 5 | Human Fit (PortaCount) | Stanford University | 5 | 5 | 5 |
| 4C Air Inc. | MBL 5x | Human Fit (PortaCount) | Stanford University | 5 | 5 | 5 | Human Fit (PortaCount) | Stanford University | 5 | 5 | 5 |
| 4C Air Inc. | DH 5x | Human Fit (PortaCount) | Stanford University | 5 | 5 | 5 | Human Fit (PortaCount) | Stanford University | 5 | 5 | 5 |
| Stryker Ltd. | VHP+$O_3$ 5x | Human Fit (PortaCount) | Stanford University | 5 | 5 | 5 | Human Fit (PortaCount) | Stanford University | 5 | 5 | N/A |
| | | | **Integrity Subtotal:** | **101** | **101** | **101** | | | **172** | **172** | **144** |
| | | | **GRAND TOTAL** | **128** | **128** | **158** | | | **227** | **199** | **171** |
