## Supplemental Table S2A_2B for "Addressing Personal Protective Equipment (PPE) Decontamination: Methylene Blue and Light Inactivates SARS-CoV-2 on N95 Respirators and Masks with Maintenance of Integrity and Fit"

### Table S2A: Integrity Results for Face Masks (MMs and CM)

| Integrity Test | Decontamination method | WHO Type II mask (FW) | | | Halyard type IIR mask (FH) | | | Community mask (CM) | | |
|---|---|---|---|---|---|---|---|---|---|---|
| | | N | Mean (SD) | p-value* | N | Mean (SD) | p-value* | N | Mean (SD) | p-value* |
| NaCl Filtration Efficiency (%) *(Standard: ≥95% for respirators)* | Untreated | 5 | 76.32 (1.54) | ref | 5 | 85.78 (1.71) | ref | 5 | 33.92 (13.66) | ref |
| | MBL 5x | 5 | 85.30 (0.89) | 0.012 | 5 | 93.58 (0.68) | 0.012 | 5 | 32.42 (1.91) | 0.14 |
| | DH 5x | 5 | 80.70 (4.79) | 0.21 | 5 | 86.94 (0.66) | 0.35 | 5 | 46.94 (12.65) | 0.09 |
| | VHP+$O_3$ 5x | 5 | 85.80 (1.48) | 0.012 | 5 | 93.13 (0.48) | 0.012 | N/A | N/A | N/A |
| Bacterial Filtration Efficiency (%) *(Standard: ≥98% for EN 14683 and 98≥% ASTM F2100)* | Untreated | 10 | 99.38 (0.20) | ref | 10 | 99.72 (0.16) | ref | 10 | 87.59 (4.06) | ref |
| | MBL 5x | 10 | 99.40 (0.26) | 0.85 | 10 | 99.74 (0.14) | 0.88 | 10 | 90.88 (2.40) | 0.04 |
| | DH 5x | 10 | 99.23 (0.14) | 0.07 | 10 | 99.81 (0.10) | 0.18 | 10 | 86.59 (2.21) | 0.50 |
| | VHP+$O_3$ 5x | 5 | 99.20 (0.16) | 0.10 | 5 | 99.64 (0.13) | 0.31 | N/A | N/A | N/A |
| Breathing Resistance with NaCl Inhalation mm$H_2$0 *(Standard: ≤35 mmH2O for respirators)* | Untreated | 3 | 9.37 (0.74) | ref | 3 | 11.87 (2.05) | ref | 3 | 8.10 (1.42) | ref |
| | MBL 5x | 3 | 8.97 (0.81) | 0.56 | 3 | 9.50 (1.73) | 0.20 | 3 | 4.20 (0.00) | 0.04 |
| | DH 5x | 3 | 8.93 (0.90) | 0.55 | 3 | 8.43 (1.91) | 0.10 | 3 | 3.43 (1.42) | 0.02 |
| | VHP+$O_3$ 5x | 3 | 8.43 (1.50) | 0.39 | 3 | 10.33 (0.81) | 0.29 | N/A | N/A | N/A |
| Breathing Resistance with NaCl Exhalation mm$H_2$0 *(Standard: ≤25 mm H2O for respirators)* | Untreated | 3 | 3.30 (0.17) | ref | 3 | 4.13 (0.15) | ref | 3 | 4.70 (0.69) | ref |
| | MBL 5x | 3 | 3.97 (0.06) | 0.003 | 3 | 5.33 (0.12) | 0.0004 | 3 | 3.60 (0.10) | 0.11 |
| | DH 5x | 3 | 3.83 (0.25) | 0.04 | 3 | 4.80 (0.40) | 0.054 | 3 | 3.50 (0.61) | 0.09 |
| | VHP+$O_3$ 5x | 3 | 4.17 (0.38) | 0.02 | 3 | 5.60 (0.26) | 0.001 | N/A | N/A | N/A |
| Pressure Drop with Bacterial Filtration Efficiency *(Standard: EN 14683:2019 Annex C <40 Pa/cm2 for Type II and ASTM F2100 spec <6.0 mm H2O/cm2 [58.83 Pa/cm2 for Type IIR)* | Untreated | 10 | 29.09 (1.76) | ref | 10 | 40.98 (1.03) | ref | 10 | 48.99 (2.17) | ref |
| | MBL 5x | 10 | 29.09 (1.66) | 0.99 | 10 | 41.16 (2.19) | 0.82 | 10 | 51.71 (4.41) | 0.10 |
| | DH 5x | 10 | 27.78 (1.69) | 0.10 | 10 | 40.03 (2.54) | 0.29 | 10 | 51.22 (3.98) | 0.14 |
| | VHP+$O_3$ 5x | 5 | 32.30 (1.70) | 0.005 | 5 | 44.48 (3.30) | 0.08 | N/A | N/A | N/A |
| Sheffield dummy fit inhalation 28.3 l/m *No min requirements for face masks (Modified version EN 149 7.16 was used; Maximum permitted resistance (mbar), inhalation: 0.7 (30/l/min); 2.4 (95 l/min) ) exhalation (160 l/min 3.0)* | Untreated | 3 | -0.18 (0.01) | ref | 3 | -0.16 (0.02) | ref | 3 | -0.15 (0.02) | ref |
| | MBL 5x | 3 | -0.19 (0.01) | 0.26 | 3 | -0.16 (0.01) | 0.82 | 3 | -0.15 (0.02) | 0.80 |
| | DH 5x | 3 | -0.17 (0.01) | 0.99 | 3 | -0.14 (0.01) | 0.48 | 3 | -0.17 (0.02) | 0.41 |
| | VHP+$O_3$ 5x | 3 | -0.18 (0.02) | 0.99 | 3 | -0.15 (0.01) | 0.64 | N/A | N/A | N/A |
| Sheffield dummy fit inhalation 85 l/m | Untreated | 3 | -0.54 (0.06) | ref | 3 | -0.43 (0.05) | ref | 3 | -0.49 (0.03) | ref |
| | MBL 5x | 3 | -0.61 (0.04) | 0.17 | 3 | -0.47 (0.01) | 0.31 | 3 | -0.48 (0.03) | 0.72 |
| | DH 5x | 3 | -0.55 (0.02) | 0.78 | 3 | -0.46 (0.02) | 0.40 | 3 | -0.56 (0.08) | 0.20 |
| | VHP+$O_3$ 5x | 3 | -0.56 (0.07) | 0.72 | 3 | -0.44 (0.03) | 0.66 | N/A | N/A | N/A |

| | | | | | | | | | | |
|---|---|---|---|---|---|---|---|---|---|---|
| Sheffield dummy fit exhalation 85 l/m | Untreated | 3 | 0.51 (0.05) | ref | 3 | 0.37 (0.04) | ref | 3 | 0.47 (0.03) | ref |
| | MBL 5x | 3 | 0.55 (0.03) | 0.22 | 3 | 0.42 (0.01) | 0.13 | 3 | 0.47 (0.04) | 0.92 |
| | DH 5x | 3 | 0.51 (0.01) | 0.82 | 3 | 0.39 (0.03) | 0.51 | 3 | 0.53 (0.05) | 0.16 |
| | VHP+$O_3$ 5x | 3 | 0.52 (0.03) | 0.63 | 3 | 0.40 (0.02) | 0.27 | N/A | N/A | N/A |
| Sheffield dummy fit exhalation 160 l/m | Untreated | 3 | 0.97 (0.02) | ref | 3 | 0.71 (0.06) | ref | 3 | 0.92 (0.05) | ref |
| | MBL 5x | 3 | 1.05 (0.04) | 0.37 | 3 | 0.81 (0.02) | 0.07 | 3 | 0.95 (0.09) | 0.63 |
| | DH 5x | 3 | 0.97 (0.02) | 0.85 | 3 | 0.73 (0.03) | 0.70 | 3 | 1.06 (0.13) | 0.16 |
| | VHP+$O_3$ 5x | 3 | 1.02 (0.08) | 0.68 | 3 | 0.76 (0.04) | 0.33 | N/A | N/A | N/A |
| Fluid Penetration inside surface (% pass)[1] *(no Standard)* | Untreated | 10 | 60% | ref | 10 | 90% | ref | 10 | 30% | ref |
| | MBL 5x | 10 | 80% | 0.63 | 10 | 100% | 0.99 | 10 | 40% | 0.99 |
| | DH 5x | 10 | 50% | 0.99 | 10 | 90% | 0.99 | 10 | 50% | 0.65 |
| | VHP+$O_3$ 5x | 5 | 40% | 0.61 | 5 | 100% | 0.99 | N/A | N/A | N/A |
| Fluid Penetration outside surface (% pass)[6] *(Standard: ≥90.625% should pass: 29 out of 32 samples according to ASTM F2100)* | Untreated | 15 | 33.3% | ref | 15 | 40% | ref | 15 | 93.3% | ref |
| | MBL 5x | 10 | 50% | 0.44 | 10 | 40% | 0.99 | 10 | 60% | 0.12 |
| | DH 5x | 10 | 50% | 0.44 | 10 | 50% | 0.70 | 10 | 70% | 0.27 |
| | VHP+$O_3$ 5x | 5 | 100% | 0.03 | 5 | 100% | 0.04 | N/A | N/A | N/A |
| Human fit (PortaCount) Overall Fit Factor *(No minimum requirements for face masks. Standard for Respirators: Fit factor≥100; OSHA 29 CFR 1910.134(f))* | Untreated | 5 | 3.81 (0.82) | ref | 5 | 3.72 (1.00) | ref | 5 | 3.34 (0.78) | ref |
| | MBL 5x | 5 | 4.11 (0.83) | 0.58 | 5 | 3.54 (0.67) | 0.99 | 5 | 2.94 (1.01) | 0.40 |
| | DH 5x | 5 | 3.63 (0.87) | 74 | 5 | 3.65 (1.72) | 0.53 | 5 | 2.76 (0.67) | 0.30 |
| | VHP+$O_3$ 5x | 5 | 2.41 (0.26) | 0.02 | 5 | 2.17 (0.39) | 0.012 | N/A | N/A | N/A |
| Tensile strength force (Newtons) *(no Standard)* | Untreated | N/A | N/A | N/A | N/A | N/A | N/A | 6 | 194.35 (3.58) | ref |
| | MBL 5x | N/A | N/A | N/A | N/A | N/A | N/A | 15 | 170.7 (20.53) | 0.006 |
| | DH 5x | N/A | N/A | N/A | N/A | N/A | N/A | 15 | 206.41 (25.62) | 0.08 |
| Tensile strength: Elongation (%) *(no Standard)* | Untreated | N/A | N/A | N/A | N/A | N/A | N/A | 6 | 11.13 (0.21) | ref |
| | MBL 5x | N/A | N/A | N/A | N/A | N/A | N/A | 15 | 30.35 (5.78) | <0.0001 |
| | DH 5x | N/A | N/A | N/A | N/A | N/A | N/A | 15 | 15.33 (9.87) | 0.90 |
| Tensile strength: Maximum load (190% strain) *(no Standard)* | Untreated | 12 | 0.44 (0.05) | ref | 12 | 0.44 (0.04) | ref | N/A | N/A | N/A |
| | MBL 5x | 25 | 0.46 (0.03) | 0.21 | 25 | 0.39 (0.04) | 0.001 | N/A | N/A | N/A |
| | DH 5x | 21 | 0.41 90.03) | 0.08 | 21 | 0.34 (0.05) | 0.0001 | N/A | N/A | N/A |
| | VHP+$O_3$ 5x | 10 | 0.34 (0.03) | <0.0001 | 10 | 0.35 (0.03) | <0.0001 | N/A | N/A | N/A |

** Difference between results from decontaminated face masks compared to untreated masks (Student's t test, Mann-Whitney U test or Fisher's exact test). Ref = reference category*

**Table 2B: Integrity Testing Results for Respirators (FFRs)**

| Integrity Test | Decontamination method | Halyard Duck Bill Respirator (RH) | | | 3M half sphere Respirator (RM) | | | 3M 1870+ panel Respirator (R3) | | |
|---|---|---|---|---|---|---|---|---|---|---|
| | | N | Mean (SD) | p-value | N | Mean (SD) | p-value | N | Mean (SD) | p-value |
| NaCl filtration efficiency (%) *(≥95%)* | Untreated | 4 | 99.22 (0.17) | ref | 4 | 98.73 (0.65) | ref | 2 | 99.13 (0.41) | ref |
| | MBL 5x | 7 | 98.98 (0.78) | 0.78 | 7 | 98.87 (0.24) | 0.92 | 7 | 99.78 (0.09) | 0.057 |
| | DH 5x | 7 | 99.22 (0.28) | 0.78 | 7 | 98.79 (0.24) | 0.78 | 7 | 99.39 (0.49) | 0.46 |
| | VHP+$O_3$ 5x | 7 | 98.92 (0.58) | 0.78 | 7 | 98.47 (0.40) | 0.22 | 7 | 99.72 (0.15) | 0.057 |
| NaCl filter efficiency (% after loading) | Untreated | 3 | 99.07 (0.33) | ref | 3 | 98.96 (0.27) | ref | 5 | 98.57 (1.75) | ref |
| | MBL 5x | 3 | 98.72 (0.53) | 0.39 | 3 | 99.21 (0.13) | 0.22 | 3 | 98.92 (1.34) | 0.99 |
| | DH 5x | 3 | 98.77 (0.50) | 0.44 | 3 | 98.65 (0.30) | 0.25 | 3 | 99.23 (0.78) | 0.99 |
| | VHP+$O_3$ 5x | 3 | 99.12 (0.26) | 0.82 | 3 | 98.75 (0.22) | 0.37 | 3 | 99.56 (0.16) | 0.99 |
| Breathing Resistance with NaCl Inhalation[1] *(≤35 mm$H_2O$)* | Untreated | 3 | 8.81 (0.14) | ref | 3 | 7.87 (0.00) | ref | 5 | 5.33 (0.78) | ref |
| | MBL 5x | 3 | 8.38 (0.00) | 0.04 | 3 | 7.20 (0.39) | 0.06 | 3 | 6.86 (0.26) | 0.02 |
| | DH 5x | 3 | 8.30 (0.14) | 0.012 | 3 | 6.69 (0.15) | 0.06 | 3 | 6.26 (0.39) | 0.11 |
| | VHP+$O_3$ 5x | 3 | 8.21 (0.39) | 0.07 | 3 | 6.77 (0.15) | 0.06 | 3 | 6.35 (0.44) | 0.09 |
| Breathing Resistance with NaCl Exhalation[2] *(≤25 mm$H_2O$)* | Untreated | 3 | 8.89 (0.00) | ref | 3 | 7.79 (0.14) | ref | 5 | 4.88 (0.58) | ref |
| | MBL 5x | 3 | 8.38 (0.00) | <0.0001 | 3 | 7.03 (0.39) | 0.03 | 3 | 6.10 (0.26) | 0.015 |
| | DH 5x | 3 | 8.21 (0.14) | 0.015 | 3 | 6.77 (0.29) | 0.006 | 3 | 6.18 (0.39) | 0.014 |
| | VHP+$O_3$ 5x | 3 | 8.72 (0.39) | 0.53 | 3 | 6.86 (0.00) | 0.008 | 3 | 5.76 (0.29) | 0.053 |
| Sheffield dummy fit inhalation 28.3 l/m *(Modified version EN 149 7.16 was used; Maximum permitted resistance (mbar), inhalation: 0.7 (30/l/min); 2.4 (95 l/min) ) exhalation (160 l/min 3.0)* | Untreated | 3 | -0.27 (0.01) | ref | 3 | -0.26 (0.02) | ref | 3 | -0.22 (0.01) | ref |
| | MBL 5x | 3 | -0.29 (0.02) | 0.20 | 3 | -0.26 (0.01) | 0.99 | 3 | -0.23 (0.01) | 0.52 |
| | DH 5x | 3 | -0.26 (0.02) | 0.48 | 3 | -0.21 (0.01) | 0.007 | 3 | -0.25 (0.01) | 0.008 |
| | VHP+$O_3$ 5x | 3 | -0.29 (0.02) | 0.28 | 3 | -0.25 (0.02) | 0.37 | 3 | -0.22 (0.01) | 0.23 |
| Sheffield dummy fit inhalation 85 l/m | Untreated | 3 | -0.85 (0.03) | ref | 3 | -0.79 (0.05) | ref | 3 | -0.66 (0.03) | ref |
| | MBL 5x | 3 | -0.88 (0.04) | 0.36 | 3 | -0.78 (0.04) | 0.79 | 3 | -0.68 (0.03) | 0.41 |

| | | | | | | | | | | |
|---|---|---|---|---|---|---|---|---|---|---|
| | DH 5x | 3 | -0.80 (0.06) | 0.27 | 3 | -0.61 (0.01) | 0.004 | 3 | -0.75 (0.03) | 0.014 |
| | VHP+$O_3$ 5x | 3 | -0.87 (0.04) | 0.58 | 3 | -0.74 (0.04) | 0.27 | 3 | -0.65 (0.01) | 0.57 |
| Sheffield dummy fit exhalation 85 l/m | Untreated | 3 | 0.80 (0.03) | ref | 3 | 0.74 (0.06) | ref | 3 | 0.63 (0.04) | ref |
| | MBL 5x | 3 | 0.83 (0.03) | 0.26 | 3 | 0.75 (0.03) | 0.93 | 3 | 0.66 (0.02) | 0.48 |
| | DH 5x | 3 | 0.78 (0.02) | 0.30 | 3 | 0.59 (0.01) | 0.04 | 3 | 0.73 (0.01) | 0.008 |
| | VHP+$O_3$ 5x | 3 | 0.84 (0.04) | 0.26 | 3 | 0.72 (0.03) | 0.61 | 3 | 0.62 (0.01) | 0.88 |
| Sheffield dummy fit exhalation 160 l/m | Untreated | 3 | 1.62 (0.05) | ref | 3 | 1.49 (0.12) | ref | 3 | 1.24 (0.03) | ref |
| | MBL 5x | 3 | 1.66 (0.01) | 0.19 | 3 | 1.46 (0.05) | 0.99 | 3 | 1.28 (0.05) | 0.38 |
| | DH 5x | 3 | 1.61 (0.02) | 0.92 | 3 | 1.15 (0.01) | 0.08 | 3 | 1.43 (0.03) | 0.08 |
| | VHP+$O_3$ 5x | 3 | 1.69 (0.05) | 0.11 | 3 | 1.46 (0.02) | 0.99 | 3 | 1.22 (0.02) | 0.38 |
| Human fit (PortaCount) Overall Fit Factor *(Standard: Fit factor ≥100; OSHA 29 CFR 1910.134(f)* | Untreated | 10 | 150.13 (54.95) | ref | 10 | 149.53 (51.79) | ref | 10 | 175.70 (36.19) | ref |
| | MBL 5x | 10 | 170.28 (48.76) | 0.39 | 10 | 142.59 (51.26) | 0.56 | 10 | 165.51 (50.53) | 0.49 |
| | DH 5x | 10 | 173.82 (37.93) | 0.31 | 10 | 147.23 (46.95) | 0.56 | 9 | 184.42 (45.00) | 0.46 |
| | VHP+$O_3$ 5x | 10 | 91.67 (82.93) | 0.14 | 10 | 72.30 (67.33) | 0.011 | 10 | 159.41 (65.23) | 0.77 |
| Manikin (Advanced Headform) Overall Fit Factor *(Standard: ≥100 per OSHA 1910.134 (f)(7) through quantitative fit testing)* | Untreated | 3 | 188.00 (20.78) | ref | 3 | 200.00 (0.00) | ref | 3 | 195.67 (7.51) | ref |
| | MBL 5x | 2 | 186.50 (19.09) | 0.99 | 3 | 188.00 (20.78) | 0.51 | 3 | 159.33 (13.01) | 0.08 |
| | DH 5x | 3 | 163.00 (32.91) | 0.35 | 3 | 200.00 (0.00) | 0.99 | 3 | 165.67 (45.08) | 0.18 |
| | VHP+$O_3$ 5x | 3 | 186.00 (24.25) | 0.99 | 3 | 127.67 (20.78) | 0.06 | 3 | 195.67 (7.51) | 0.99 |
| Tensile strength: Force (Newtons) in top strap *(no Standard)* | Untreated | 4 | 2.41 (0.03) | ref | 4 | 2.83 (0.22) | ref | 2 | 1.71 (0.1) | ref |
| | MBL 5x | 5 | 2.37 (0.02) | 0.39 | 5 | 2.73 (0.10) | 0.71 | 5 | 1.77 (0.17) | 0.85 |
| | DH 5x | 5 | 2.31 (0.01) | 0.02 | 5 | 2.71 (0.04) | 0.90 | 5 | 1.87 (0.03) | 0.08 |
| | VHP+$O_3$ 5x | 5 | 1.99 (0.07) | 0.02 | 5 | 3.61 (0.30) | 0.02 | 5 | 1.92 (0.04) | 0.08 |
| Tensile strength: Force (Newtons) in bottom strap *(no Standard)* | Untreated | 4 | 2.46 (0.04) | ref | 4 | 2.71 (0.20) | ref | 2 | 1.71 (0.15) | ref |
| | MBL 5x | 5 | 2.42 (0.04) | 0.39 | 5 | 2.76 (0.10) | 0.54 | 5 | 1.96 (0.06) | 0.02 |
| | DH 5x | 5 | 2.33 (0.03) | 0.02 | 5 | 2.67 (0.06) | 0.99 | 5 | 1.73 (0.03) | 0.85 |
| | VHP+$O_3$ 5x | 5 | 2.00 (0.04) | 0.02 | 5 | 3.62 (0.23) | 0.02 | 5 | 1.91 (0.08) | 0.052 |

* *Difference between results from decontaminated face masks compared to untreated masks (Student's t test or Mann-Whitney U test). Ref = reference category*
