## Supplementary material for "Addressing Personal Protective Equipment (PPE) Decontamination: Methylene Blue and Light Inactivates SARS-CoV-2 on N95 Respirators and Masks with Maintenance of Integrity and Fit": DEMAND STUDY TABLE 1 Expanded

Table 1 - Virus Reduction 10μM Methylene Blue vs. 75°C Dry Heat

| | | MHV | | PRCV | | SARS-CoV-2 (Lab 1) | | SARS-CoV-2 (Lab 2) | | SARS-CoV-2 avg. | | Overall avg. | |
|---|---|---|---|---|---|---|---|---|---|---|---|---|---|
| Condition | Mask type | Log reduction | % reduction | Log reduction | % reduction | Log reduction | % reduction | Log reduction | % reduction | Log reduction | % reduction | Log reduction | % reduction |
| 10μM MB (w/ 30 min light) | RH | 4.4 | > 99.9% | 5.5 | > 99.9% | 2.2 | > 99.4% | 4.3 | > 99.9% | 3.2 | 99.6% | 4.1 | 99.8% |
| 10μM MB (w/ 30 min light) | RM | 3.5 | > 99.9% | 2.7 | 99.8% | 2.8 | > 99.8% | 4.0 | > 99.9% | 3.4 | 99.9% | 3.2 | 99.9% |
| 10μM MB (w/ 30 min light) | *R3 | 5.1 | > 99.9% | 5.0 | > 99.9% | 3.3 | > 99.9% | 3.1 | > 99.9% | 3.2 | > 99.9% | 4.1 | > 99.9% |
| 10μM MB (w/ 30 min light) | *FW | 5.8 | > 99.9% | 5.5 | > 99.9% | 3.6 | > 99.9% | 3.1 | > 99.9% | 3.4 | > 99.9% | 4.5 | > 99.9% |
| 10μM MB (w/ 30 min light) | FH | 3.8 | > 99.9% | 5.1 | > 99.9% | 3.1 | > 99.9% | 4.2 | > 99.9% | 3.6 | > 99.9% | 4.0 | > 99.9% |
| 10μM MB (w/ 30 min light) | CM | 3.2 | > 99.9% | 0.4 | 60.5% | 1.3 | > 94.6% | 2.7 | > 99.9% | 2.0 | 97.3% | 1.9 | 88.7% |
| 75°C DH (60 min) | RH | 1.6 | 97.5% | 5.4 | > 99.9% | 2.4 | > 99.6% | 2.9 | 99.8% | 2.6 | 99.7% | 3.1 | 99.2% |
| 75°C DH (60 min) | RM | 1.4 | 95.8% | 4.7 | > 99.9% | 1.1 | 91.8% | 3.4 | 99.8% | 2.2 | 95.8% | 2.6 | 96.8% |
| 75°C DH (60 min) | *R3 | 1.8 | 98.6% | 5.3 | > 99.9% | 2.3 | 99.5% | 3.7 | > 99.9% | 3.0 | 99.7% | 3.3 | 99.5% |
| 75°C DH (60 min) | *FW | 2.1 | 99.2% | 5.5 | > 99.9% | 2.0 | > 99.1% | 3.9 | > 99.9% | 3.0 | 99.5% | 3.4 | 99.5% |
| 75°C DH (60 min) | FH | 1.7 | 98.0% | 5.1 | > 99.9% | 1.0 | > 90.6% | 3.0 | 99.6% | 2.0 | 95.1% | 2.7 | 97.0% |
| 75°C DH (60 min) | CM | 3.1 | 99.9% | 5.0 | > 99.9% | 0.9 | > 88.8% | 2.0 | > 99.9% | 1.5 | 94.3% | 2.8 | 97.1% |

*Note:*

[*] Data extracted from inactivation curve (Except for PRCV, SARS-CoV-2 (Lab 2), and MHV 10μM MB)

[†] Light source for testing MB on PRCV = 12500 LUX red light; Light source for testing MB on all other virus types = 50000 LUX white light

[‡] Due to the limit of detection of the assays used to titer the virus, the highest % reduction is indicated as >99.9%

[§] RH = Halyard duckbill respirator (Fluidshield-46727), RM = 3M half-sphere respirator (1860), R3 = 3M Panel respirator (1870+), FW = Type II EN 14683 generic face mask, FH = Type IIR ASTM F2100 Level 2 Halyard face mask, CM = Community mask

[¶] MB = Methylene Blue, DH = Dry Heat
